# Exploring the Impact of the Covid-19 Pandemic on Health-Related Behaviours: A Person-Centred Analysis of Two British Longitudinal Cohort Studies

**DOI:** 10.1101/2023.03.28.23287685

**Authors:** Janet Kiri, Valerie Brandt

## Abstract

The current study explored the impact of the Covid-19 pandemic on health-related behaviours in the United Kingdom. We conducted a repeated measures latent class analysis with five indicators of health-related behaviours; frequency of alcohol consumption, binge drinking, smoking, BMI and sleep, to identify distinct subgroups of individuals with similar patterns of change across three timepoints during the first 9 months of the pandemic. We hypothesised that various psychosocial risk factors, such as a history of adverse childhood experiences would predict membership in latent classes with a higher probability of engaging in risky health behaviours, and that protective factors, like social support, would be associated with membership in classes with less risky health behaviours. We identified 5 latent classes, and multinomial logistic regression analyses revealed multiple predictors of class membership. Our findings did not support the relationship between poor mental health and the adoption of risky health behaviours.

---

The Covid-19 pandemic has been an ongoing global crisis since December 2019. In response, governments across the world imposed restrictions on social gatherings to contain the spread of the virus (Cameron-Blake et al., 2020). The pandemic and the resulting lockdown disrupted daily life, causing financial uncertainty, unemployment and social isolation (HM Government, 2020). The potential negative impact of the pandemic on people’s mental and physical health highlights the importance of exploring how individuals have coped with the stress added to their daily lives (Maric et al., 2021).

The impact of the pandemic on mental health has been a key area of interest for mental health professionals and researchers. Lockdown restrictions caused massive disruptions to patient care (Bauer-Staeb et al., 2021). Concerns were raised about the potential for these prolonged periods of isolation to increase the need for mental health services (Al-Aly, 2022). A recent meta-analysis that comprised 173 studies from 32 countries demonstrated why these concerns were warranted, with the findings suggesting that the pandemic was associated with substantial increases in the global prevalence of mental illnesses such as depression, anxiety and PTSD (Dragioti et al., 2022).

From a behavioural perspective, home confinement may have limited the amount of positive stimuli in people’s environment, reducing opportunities for positive reinforcement, and in conjunction with the constant distressing news reports of the rising death toll, lockdown may have exacerbated feelings of negative affect (Lewinsohn, 1974). Research has illustrated how a lack of positive environmental stimuli comprises a risk factor for the development of depression (Carvalho & Hopko, 2011), therefore, the lockdown restrictions may have contributed to the observed increase in mental health symptomatology worldwide (COVID-19 Mental Disorders Collaborators, 2021).

There are a range of cognitive, affective, and behavioural strategies that individuals employ when attempting to adapt and cope with negative affect. For instance, people may engage in adaptive coping strategies to mitigate negative emotionality (Brown et al., 2005). Emotion-focused coping strategies, such as exercising (Cairney et al., 2014) or seeking support from loved ones (Holahan et al., 2004) facilitate the regulation of negative emotions and mitigate the impact of stressful events on their psychological wellbeing. Problem focused coping strategies comprise an attempt to alter one’s own behaviour to address the external or internal stimulus that is eliciting negative affect (Aldwin & Yancura, 2004). However, the Covid-19 pandemic conveys unique barriers for individuals to implement adaptive coping strategies (Ogueli et al., 2021).

Maladaptive coping strategies, such as substance use, may provide temporarily relieve from negative affect but confer long term adverse health risks (Blevins et al., 2016). As postulated by Lazarus and Folkman (1984), the choice of adaptive over maladaptive coping strategies depends on whether the individual perceives the source of their stress as one that is within their power to control. The pandemic may have had a detrimental impact on people’s sense of control, referring to the belief that their actions can impact their emotional state or external environment (Langer, 1977). A review that examined how people respond to natural disasters, (Goldman & Galea, 2014), suggests that people often experience a reduced sense of control when faced with external events that incur significant changes to their lifestyle, as was the case during the early months of the Covid-19 pandemic.

In fact, a low sense of control has shown a negative association with the ability to delay gratification in experimental studies (Whitson & Galinsky, 2008). Diminished ability to delay gratification, defined as the ability to delay the receipt of an immediately available reward in favour of a long-term reward (Tobin & Graziano, 2010), has been implicated in the choice to use alcohol (Wulfert et al., 2002), nicotine (MacKillop et al., 2012) and the excessive consumption of unhealthy foods (Kemp et al., 2013) to cope with negative affect. Hence, the loss of control elicited by the restrictions placed on movement during lockdown, uncertainty over the virus and reduced opportunities for positive reinforcement, may result in the adoption of maladaptive coping strategies to mitigate psychological distress.

The following sections will explore the impact of the Covid-19 pandemic on health-related behaviours (HRB) such as sleep, alcohol consumption, smoking, physical activity and dietary choices.

## Alcohol Consumption

Alcohol related deaths in the UK were rising annually before the start of the pandemic (Office for National Statistics, 2021), and a recent longitudinal study by Public Health England (2021) found an increase in the proportion of people who reported problematic drinking behaviour during the first months of lockdown. One plausible explanation for this finding arises from the self-medication hypothesis (Khantzian, 2003) which posits that an increase in alcohol consumption is motivated by the desire to attenuate negative affect. Indeed, findings from the UK household longitudinal study indicate that at the beginning of the pandemic the proportion of adults reporting clinically significant levels of psychological distress rose by 8% compared to the previous year (National Mental Health Intelligence Network, 2022) and a recent meta-analysis that examined 128 studies from 58 countries found a positive association between psychological distress and increased alcohol consumption during the first months of the pandemic (Acuff et al., 2022), supporting the notion that more people may be drinking to cope with distress during lockdown.

On the other hand, the closure of non-essential shops and pubs during the first months of lockdown in May 2020 may have contributed to a reduction in alcohol consumption. Indeed, Villadsen et al. (2021) found that high risk drinking was lower in May 2020 compared to before the pandemic. In line with the availability hypothesis (Rabow & Watts, 1983), the decline in problematic drinking could be attributable to an inability to conduct social activities that involve alcohol, due to social distancing, and the early closure of shops during May 2020. Therefore, it is important that research examines the impact of lockdown on alcohol use behaviour.

## Smoking

Tobacco consumption has consistently been linked to an increased risk for adverse health outcomes, such as cardiovascular disease and several cancers (WHO, 2022). Official statistics suggest that the prevalence of smoking in the UK is declining each year (ONS, 2020). With the closure of shops limiting access to tobacco products and the increased vulnerability of smokers to respiratory viruses like Covid-19 (Feldman & Anderson, 2013), lockdown restrictions may result in a decline in smoking behaviour. In support of this, Jackson and colleagues (2021) conducted a cross-sectional survey in the UK that showed how smokers were more likely to attempt cessation during lockdown compared to before the pandemic.

Interestingly, in the Jackson et al. (2021) study, these cessation attempts did not correspond to a decreased prevalence of smoking, indicating that despite the motivation to quit, many attempts were not successful. In the context of the heightened negative affect many experienced during lockdown (National Mental Health Intelligence Network, 2022) the stress experienced during lockdown may be the impetus for relapse amongst those attempting to quit. Mckee et al. (2014) illustrated how psychological distress reduced smokers’ ability to delay the impulse to smoke in a lab study, suggesting that smokers may attempt to quit, but their likelihood of success may be diminished due to the stress associated with the pandemic.

## Exercise, Diet and Weight Gain

As of 2018, 63% of adults in the UK were overweight or obese (NHS, 2020) and the psychological distress associated with the pandemic may inadvertently contribute to the escalating burden of obesity on the NHS. Two factors underlie the potential acceleration of weight gain in the UK population during the pandemic; reduced physical activity due to lockdown restrictions and increased emotional eating as a coping mechanism for heightened negative affect. The impact of poor diet and physical inactivity on weight gain is well supported (Miller et al., 1997), suggesting that changes in diet and exercise may elicit weight gain during the pandemic. Indeed, the link between negative affect and increased food intake has been demonstrated in experimental studies (Cardi et al, 2015), suggesting that some may use emotional eating in response to the stress elicited during the pandemic (Macht, 2008). Emotional eating refers to overeating as a coping strategy in response to negative emotionality (Frayn & Knäuper, 2018) and recent research conducted during the pandemic suggests that those experiencing high levels of psychological distress were more likely to engage in emotional eating (Shen et al., 2020). Furthermore, emotional eaters are more likely to consume foods high in calories, fat, sodium and sugar (Camilleri et al., 2014), increasing the risk of weight gain (Vandevijvere et al., 2014).

The impact of the pandemic on activity levels is less clear. Kass and colleagues (2021) examined activity levels during the first lockdown in England and showed that while exercising at home and at the gym decreased considerably, more people engaging in outdoor forms of exercise. On the other hand, Dicken et al. (2020) found significant increases in the BMI of adults across the UK suggesting that regardless of increased outdoor exercise, the pandemic had an impact on weight gain that requires further exploration (NHS, 2019).

## Sleep

Sleep is crucial for resilience against psychological distress (Hughes et al., 2018) and immunological function (Besedovsky et al., 2012). During lockdown, home confinement and school closures necessitated that people adapt to managing increased parental responsibilities, acclimatise to working from home and protect themselves from the virus (Altena et al., 2020). These lifestyle disruptions and reduced opportunities for physical activity may have had a detrimental impact on people’s quality of sleep (Chennaoui et al., 2015). Studies suggest that emotional distress may have a reciprocal relationship with sleep, as high levels of distress disrupt the ability to sleep, increasing daytime tiredness, impairing emotional regulatory skills (Vandekerckhove & Wang, 2017) and further exacerbating feelings of negative affect (Zhang et al., 2022). Hence, the protective nature of sleep for both mental and physical health highlights the importance of examining the impact of the pandemic on sleep.

## Adverse Childhood Experiences

A population who may be more susceptible to experiencing mental health difficulties comprises adults with a history of adverse childhood experiences (ACE) (McCroy et al., 2017). According to the theory of latent vulnerability, exposure to ACEs results in neurocognitive alterations that confer short term benefits in the adverse environment but may increase the risk of developing a mental illness when exposed to stress during adulthood and adolescence, depending on the individual and/or familial factors that provide resilience or increase the risk of developing mental illness (McCroy & Viding, 2015). Prior research has shown that individuals with a history of ACEs show disrupted emotion regulation and heightened feelings of negative affect in response to stress, suggesting that this population may be more susceptible to engaging in maladaptive health behaviours, like drinking, smoking and overeating in order to alleviate psychological distress (McLaughlin et al., 2015; Sacco et al., 2017; Mahmood et al., 2022). Additionally, Sullivan et al. (2019) found that ACEs were associated with chronic sleep shortages, suggesting that the neuroadaptations resulting from ACEs also have a detrimental impact on sleep. Therefore, adults with a history of ACEs may have an increased susceptibility to adverse health outcomes during the pandemic, highlighting the importance of research to examine the impact of the pandemic on this population.

## Co-occurring Health Behaviours

HRBs like alcohol use, diet, exercise, smoking and sleep have been found to co-occur within individuals (Spring et al., 2012). For instance, Schoenborn and Adams (2008) found a higher prevalence of smoking and drinking more than five drinks in a session amongst participants who reported sleeping less than seven hours a night, compared to those who slept seven to eight hours, suggesting that risky HRBs tend to co-occur with one another. Considering the increased risk of negative health outcomes associated with individual risky HRBs, the cumulative effect of multiple may convey even worse outcomes (Appleton et al., 2004).

Additionally, interventions that address a single HRB, fail to address the negative health outcomes posed by co-occurring risky HRBs, suggesting that interventions targeting multiple HRBs may be more cost-effective and efficacious for improving overall health outcomes. Further research is needed to inform such approaches through identifying subgroups of people who engaging in similar HRBs during the pandemic as this could benefit preventative interventions and facilitate the identification of psychosocial factors that could be targeted for intervention within similar clinical populations after the pandemic.

## Current Study

As individuals likely responded differently to the COVID-19 pandemic, exploring changes in HRBs may be benefitted by a person-centred approach. As opposed to variable centred methods, such as logistic regression which allow investigation into the differing factors associated with the adoption of HRBs, person-centred methods like latent class analyses facilitate the identification of subgroups of people with similar HRBs, and their adoption of HRBs is assumed to be driven by their membership in a latent class (Eye & Weiderman, 2015). Latent variable methods have gained increased interest in the behavioural change literature (Fitzpatrick et al., 2015), due to their ability to preserve heterogeneity in the data and provide a data driven method to identify naturally occurring subgroups of people within the data (Qiu et al., 2018). Additionally, as latent variable models are measurement models, multiple indicators of HRBs can be entered into the model simultaneously, reducing the likelihood of type 1 error by eliminating the need to analyse each health behaviour independently (Lanza & Rhoades, 2013).

The current study used a repeated measures latent class analysis (RMLCA) to explore whether qualitatively different and distinct subgroups of individuals who share similar trajectories in their alcohol consumption, weight gain, sleep duration and smoking could be identified^1^. In addition, we aimed to investigate whether risk factors, such as exposure to adverse life events during the pandemic, loneliness, psychological distress, mental health symptomatology and a history of ACEs as well as protective factors, like social support and social contact were predictive of class membership.

The purpose of the study was exploratory, and the data-driven nature of latent variable models rejects the imposition of any a priori hypotheses regarding the number of latent classes that may be identified, hence no such hypotheses were made (Qiu et al, 2018). However, we anticipated that some classes would demonstrate a high likelihood of engaging in several co-occurring risky HRBs, whilst others may have relatively low likelihood of engaging in any. Furthermore, we hypothesised that the high levels of loneliness, experiencing adverse life events during the pandemic, a history of adverse childhood experiences, poor mental health, a lack of social support and low social contact would be predictive of membership in a latent class with a high likelihood of engaging in risky HRBs.

## Method

### Design

The current study utilises a within-subjects design to examine patterns of five HRBs over three timepoints during the Covid-19 pandemic. The datasets were obtained from the UK data service and ethical approval was received from the University of Southampton Ethics Committee (ERGO: 62740.A2). A repeated measures latent class analysis (RMLCA) was conducted to identify distinct qualitatively different, and mutually exclusive, subgroups of individuals based on their pattern of responses to the five HRB questionnaire items at each of the three timepoints (Collins & Lanza, 2010).

### Sample

The sample data was obtained from two national longitudinal cohort studies, the Millennium cohort study (MCS) and the 1970 British Cohort Study (BCS70).

The MCS sample comprises both cohort members and their parents from across the UK who have been followed since the cohort member’s birth in 2000-2001 (Joshi & Fitzsimons, 2016). The initial sample had 18,818 participants. Prior to the Covid-19 surveys, cohort members and parents took part in seven waves conducted at regular intervals throughout the child’s life (9 months, 3 years, 5 years, 7 years, 11 years, 14 years and 17 years). These surveys enquired about various aspects of the cohort members and their parent’s lives, including mental health, their financial situation and the parent-child relationship. The MCS has a complex sample design to ensure that underrepresented groups are properly represented and incorporates stratification and clustering based on electoral wards to over-represent regions with high prevalence of ethnic minorities and child poverty (Centre for Longitudinal Studies, 2010).

The BCS70 cohort comprised an initial sample of 17,196 cohort members born in the UK during a single week in 1970 (Elliott & Shepherd, 2006). Cohort members and their parents took part in 10 waves prior to the Covid-19 surveys and provided information regarding the child’s development at regular intervals, from birth up until the age of 46 (Sullivan et al., 2022).

During the Covid-19 pandemic in 2020/2021, cohort members completed three surveys administered by the Centre for longitudinal studies which enquired about a wide range of topics, including the impact of the pandemic on their mental health, health behaviours and social lives.

**Figure 1.**
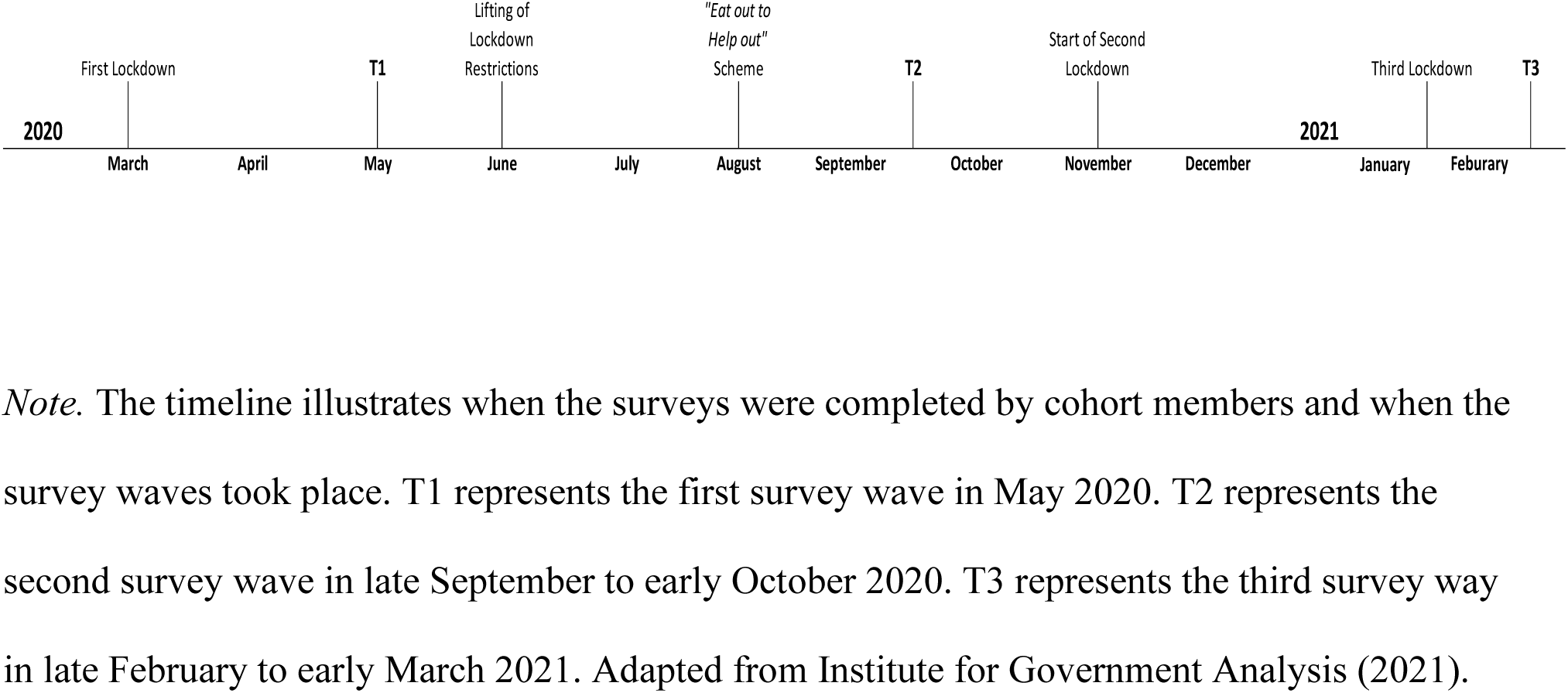
Timeline of Key Dates.

The Covid-19 surveys were originally issued to 26,036 participants from the MCS and BCS70 cohorts, out of the 10,232 participants who took part in all three online survey waves, 2,847 participants were included in the final analysis as they provided sufficient information regarding health behaviours, history of adverse childhood experiences and the other covariates of interest (Kantar Public, 2021). The demographic information for both cohorts collected during the first wave of the survey is presented in Table 1.

**Table 1.** Descriptive Statistics of Baseline Characteristics.

| Variable | BCS70 |  | MCS |  | Full Sample |  |
| --- | --- | --- | --- | --- | --- | --- |
|  | n | % | n | % | n | % |
| Gender |  |  |  |  |  |  |
| Male | 610 | 36.33 | 306 | 26.20 | 916 | 32.17 |
| Female | 1069 | 63.67 | 862 | 73.80 | 1931 | 67.83 |
| NVQ<sup>a</sup> |  |  |  |  |  |  |
| NVQ0 | 1146 | 72.58 | 1 | .13 | 1146 | 48.77 |
| NVQ1 | 175 | 11.08 | 14 | 1.81 | 189 | 8.04 |
| NVQ2 | 72 | 4.56 | 105 | 13.60 | 177 | 7.53 |
| NVQ3 | 48 | 3.04 | 95 | 12.31 | 143 | 6.09 |
| NVQ4 | 108 | 6.84 | 311 | 40.28 | 419 | 17.83 |
| NVQ5 | 30 | 1.90 | 246 | 31.87 | 276 | 11.74 |
| Ethnicity |  |  |  |  |  |  |
| White | 1660 | 98.81 | 1044 | 89.23 | 2704 | 94.88 |
| Indian | 6 | 0.36 | 20 | 1.71 | 26 | .91 |
| Mixed Race | 6 | 0.36 | 47 | 4.02 | 53 | 1.86 |
| Pakistani | 1 | 0.06 | 20 | 1.71 | 21 | .74 |
| Black African | 0 | 0.00 | 13 | 1.11 | 13 | .46 |
| Bangladeshi | 0 | 0.00 | 12 | 1.03 | 12 | .42 |
| Black Caribbean | 0 | 0.00 | 3 | .26 | 3 | .11 |
| Black Caribbean | 0 | 0.00 | 3 | .26 | 3 | .11 |
| Other Ethnic | 7 | .42 | 8 | .68 | 15 | .53 |
| Group |  |  |  |  |  |  |
| Black Caribbean | 0 | 0.00 | 3 | .26 | 3 | .11 |
| Other Ethnic | 7 | .42 | 8 | .68 | 15 | .53 |
| Group |  |  |  |  |  |  |

*Note.* $N = 2847$ . Average age was 37.4 ( $SD = 15.2$ ). Responses reflect cohort members answering "yes" to this question.
<sup>a</sup>Missing information for 497 Cohort members.

Sample size determination in latent variable models is a highly debated topic, however, some researchers have recommended sample sizes of at least 300 to detect significant effects and avoid model underidentification (Nylund-Gibson & Choi 2018). Additionally, to ensure the current study has sufficient power, a power analysis for the optimal sample size, based on the formula used for structural equation models (SEM), was also calculated as latent class analyses are considered a subset of SEM (Weller, et al., 2020). As the preliminary analysis involves testing one to nine class solutions, an a priori power analysis was computed to reflect the sample size required to detect nine latent classes (Soper, 2022). The results indicated that a sample size of 489 is needed to achieve a large effect size of .80 and an alpha level of .05 in the RMLCA (Cohen, 2013). Hence, the current sample size of 2,847 is sufficient to avoid model underidentification and test the study’s hypotheses.

### Measures

#### Adverse childhood experiences (ACEs)

The choice of measures for ACEs was operationalised in line with the ACE framework (Felitti et al., 1998) which proposes ten categories of ACEs demonstrated to be associated with poor mental and physical health outcomes. These categories comprise three types of adversity: abuse, neglect and family dysfunction. As neither cohort was asked specifically about child abuse or neglect, we included measures of victimisation that cohort members experienced before the age of 16. Experiences of victimisation comprised physical, verbal and sexual assault and was included to captured out-of-home experiences of child adversity (Karamanos et al., 2022). Additionally, parenting style was also measured as previous research into ACEs has studied this with these cohorts (Wood et al., 2019). Therefore, across both cohorts, 12 indicators of adverse childhood experiences were measured: Parental Separation, Parental Death, Suboptimal Parenting Style, Exposure to Domestic Violence (MCS), Foster care (BCS70), Parental Substance Use, Incarceration, Poor Parental Mental Health, Physical Punishment and Victimisation (sexual, physical and verbal assault).

Available information regarding early adversities was collected via self-reports from cohort members and their parents. For each category of ACE, binary measures were created to represent either no exposure (0) or exposure (1) to that form of adversity before the age of 16. In line with previous research that investigated the influence of childhood adversity using the MCS and BCS70 cohorts (Bevilacqua et al., 2021; Lacey et al., 2014; Morgan et al., 2012), the ACE scores were summed to create a cumulative score with a range of 0 to 11 and formed four categories; 0 ACEs, 1 ACE and 2 ACEs and 3+ ACEs. A full description of the measures of adversity used and the ages at which they were collected in each cohort can be found in Appendix B.

#### Mental Health

Mental health was assessed through two dimensions, psychological distress and symptoms of anxiety and depression. Both psychological constructs were measured as psychological distress captures general feelings of emotional suffering that extends beyond depressive and anxious symptomatology (Viertiö et al., 2021). Additionally, psychological distress has been shown in research to mediate the association between childhood adversity and later sleep disturbances (McPhie et al., 2014) supporting the decision to measure both dimensions of mental health.

##### Psychological Distress

Psychological distress was assessed both years before the start of the pandemic, and at each timepoint using different scales for each cohort. Prior to the pandemic, psychological distress was assessed in the BCS70 cohort (age 46) using 9 items from the Malaise Inventory (Rutter et al., 1981) which is a self-report questionnaire that assesses the presence of emotional and somatic symptoms associated with depression. The items were summed to create a total score ranging from 0-9 with scores of 4 and above indicating the prescience of depressive symptomatology. Psychological distress in the MCS cohort (age 17) was measured with the six item Kessler (K6) measure that enquires about the frequency of experiencing feelings associated with depression (Kessler et al., 2003). The six items were summed to give a total score ranging from 0-24, with scores of 13 and above indicating a greater likelihood of experiencing psychological distress. The same measures for each cohort were used during each wave of the survey to assess psychological distress throughout the pandemic. Both the K6 and the Malaise Inventory demonstrate acceptable internal consistency and good construct validity in the general population (Rodger et al., 2009; Khan et al., 2014).

##### Anxious and Depressive Symptomatology

During the pandemic, the Patient Health Questionnaire (PHQ-2) and the Generalised Anxiety Disorder scale (GAD-2) was used for both cohorts to measure depressive and anxiety symptoms (Kroenke et al., 2003; 2007). The PHQ-2 is a brief screening tools for major depressive disorder and assesses the frequency of depressed mood and anhedonia. The two-item scale had a range of 0-6 with scores of 3 and above indicating high levels of depressive symptoms. The GAD-2 is a brief screening tool for symptoms of anxiety and comprises two items. The two items were summed to compile a total score ranging from 0-6 with scores of 3 and above indicating high levels of anxiety symptoms. These two scales were combined to form a unitary measure of depressive and anxiety symptoms, thus, scores ranged from 0-12 with scores of 6 and above indicating high levels of depression and anxiety symptomatology. Both the GAD-2 and PHQ-2 have demonstrated good construct validity and internal consistency in the general population, supporting the use of these measures in the current study (Lowe et al., 2010).

#### Social Support

Four aspects of social support were assessed at each timepoint; instrumental and emotional social support, social contact and perceived loneliness (Morelli et al., 2015). The same measures were used during each wave of the survey and are relevant to both cohorts. Despite methodological concerns regarding the use of single item measures for social contact, instrumental and emotional social support, they were included in the analysis to provide a more comprehensive understanding of how differing dimensions of social support influence HRBs. Additionally, the reliability and validity of single item measures has been acceptable when the item is measuring relatively unidimensional constructs, as is the case in the current study (Bowling, 2005).

##### Instrumental and Emotional Social Support

Instrumental support, defined as the practical services provided by members of one’s support network, was measured using a single questionnaire item ‘if you were sick in bed, how much could you count on the people around you to help?”. Emotional support, referring to having someone to confide in about one’s problems, was measured using a single questionnaire item “if you needed to talk about your problems and private feelings, how much would the people around you be willing to listen?” Responses for both questions were on a Likert scale (0 = *not at all* to 4 = *a great deal),* both items were dichotomised to create two binary variables, with 0 (*not at all)* and 1 (*a little*) collapsed into one category indicating 0 (*low levels of support)*, and the last two response options were collapsed to represent 1 (*high levels of support)*.

##### Social Contact

The degree of social contact was measured with three questionnaire items that asked how many days in the past week cohort members had contacted a non-household member either “in person”, “via phone or video call” and “by email, text or other electronic messaging”. Additionally, two questionnaire items asked about the number of days in the last week cohort members took part in an online community activity and gave assistance to a non-household member affected by lockdown restrictions. Responses were rated as (0 = *never*, 1 = *one day*, 2 = *2-3 days*, 3 = *4-6 da*ys) and (4 = *everyday*). A binary variable was creased to reflect either (0) *low levels of social contact*, referring to contact with non-household members less than 4 days a week, or (1) *high levels of social contact*, referring to 4 or more days a week of non-household social contact.

##### Perceived Loneliness

The UCLA Loneliness Scale was used to measure the frequency of feelings of loneliness (Russell, 1996). Four out of the original 20 items were included in the survey and asked about how often cohort members feel that they felt “left out”, “isolated from others”, “lonely” and whether they felt that they “lack companionship”. Response options include rating the frequency of these feelings as 0 (*hardly ever*), 1 (*some of the time*), and 2 (*often*). The four items were summed to form a total score ranging from 0-8, with higher scores indicating higher levels of loneliness. Prior research indicates that the full 20 item questionnaire has high internal reliability and good test-retest reliability in samples diverse in age and ethnicity, supporting the use of this measure to assess within-subject changes in loneliness over a 9-month period (Russell, 1996).

#### Adverse Life Events

Participants were asked at T2 and T3 whether since T1, they had experienced any adverse life events, such as the death of loved ones, marital conflict, loss of employment, homelessness, serious injury and victimisation. A binary variable was created, with positive responses to any of the questions inquiring about life events coded as indicating exposure to adverse life events during the pandemic.

#### Health Behaviours

Participants were asked questions regarding the five HRBs of interest at each timepoint. Responses were dichotomised to reflect HRBs in accordance with national guidelines.

##### Alcohol Consumption

Alcohol consumption was measured through two dimensions; alcohol volume and alcohol frequency, as both excessive alcohol consumption, referred to as binge drinking, and frequent alcohol consumption have been shown to have differing potential adverse effects on health and societal outcomes (Yoo et al., 2021l; Attard et al., 2021). Therefore, both dimensions of alcohol consumption were examined as this provides a more comprehensive understanding of patterns of drinking behaviour. Two questions from the 3 item alcohol use disorders test (AUDIT-C), a brief assessment measure used in clinical settings to screen for hazardous alcohol consumption, were administered at each timepoint. The full AUDIT-C demonstrates high convergent validity with other measures of alcohol dependency and has been validated for detecting problematic drinking in online surveys (Russell & Barry, 2021).

##### Alcohol Volume

Participants were asked “in the last four weeks, how many standard alcohol drinks they consume in a typical session of drinking”. Responses were provided on a 5-point Likert scale ranging from 0 (*1-2 drinks*) to *4* (*10+ drink*s). Binge drinking was defined as drinking 4 or more drinks for females, and 5 or more drinks for males, in a single drinking session (National Institute on Alcohol Abuse and Alcoholism, n.d.). Responses were dichotomised into a binary variable indicating whether an individual engaged in binge drinking within the preceding four-week period.

##### Alcohol Frequency

Participants were asked “in the last four weeks, how often have you had an alcoholic drink”. Response options were on a 5-point Likert scale ranging from 0 (*never*) to 4 (*4 or more times a week*). The National Health Service (NHS) recommends spreading alcohol consumption over 3 or more days (NHS, 2018). Therefore, responses were dichotomised such that drinking 4 or more times a week was recoded to indicate 1 (*4 or more times a week*) and the other response options were collapsed into 0 (*less than 4 times a week*).

##### Smoking

At each wave, participant’s current smoking status was obtained and subsequently used to form a binary variable that categorised participants as either 0 (*non/ex-smokers*) or 1 (*current smokers*). The former category was applicable to participants who either never smoked, or have currently ceased smoking, while the later reflects intermittent and daily smokers.

##### Sleep

Sleep duration was assessed with a single question asking about the average hours of sleep each night in the previous month. Responses were rounded to the nearest hour and were dichotomised to form a binary variable, with between 7 and 9 hours of sleep indicating 0 (*typical sleep duration*), and responses outside of that range coded as 1 (*atypical sleep duration*) (Hirshkowitz et al., 2015).

##### Body Mass Index

Cohort members body mass index (BMI) was calculated based on the individual’s weight and height using the formula provided by the World Health Organisation (2010), to provide a standardised measure of whether participants were overweight or obese. Information regarding height was collected from the previous wave and weight information was collected at each timepoint. BMI was dichotomised into a binary variable, with 1 (*high BMI*), indicative of a BMI categorised as either overweight or obese and 0 (*low BMI*), indicating that cohort members were either a healthy weight or underweight.

#### Covariates

Potential confounding factors were identified through previous research (McCroy et al., 2017) and were included in the analysis. They were collected at T1 and include the cohort members’ age, sex, ethnic group and educational level in addition to the presence of a mental health diagnosis. The National Vocational Qualifications level system was used to assess participants level of education, and a sixth category was added to reflect participants without any formal qualifications. For the MCS, many were still in education, so their parent’s highest level of education was substituted if this information was unavailable.

### Analysis

#### Primary Analysis

##### RMLCA

The RMLCA model comprised 15 indicators of HRBs, with five indicators collected at each timepoint. RMLCA assumes that individual response patterns to the HRB questionnaire items are driven by both the individual’s underlying latent class membership and measurement error (Collins & Lanza, 2010). Latent classes are considered mutually exclusive, and class membership is assumed to fully account for the relationship between the five HRBs. RMLCA employs an iterative approach to parameter estimation, with the goal of maximising the likelihood function to produce parameter estimates with the highest probability of producing the observed data (Agresti, 2003). The RMLCA employs two principal search algorithms; the expectation maximisation algorithm, which can accommodate for missing values and estimate parameters with incomplete information for the HRB indicators of the latent classes, and the Newton Raphson algorithm when the latent model includes covariates (Collins & Lanza, 2010; Do & Batzoglou, 2008; Dempster at al., 1977). The model was assumed to have converged if the convergence criterion of ≤ .000001, reflecting the degree that parameter estimates change between each iteration, was met before the algorithm performed the maximum number of iterations (Colins & Lanza, 2010).

#### Unconditional Models

First, one to nine class unconditional models were fit to the data to identify the model with the optimal fit based on the G^2^ likelihood ratio test statistic, information criteria and the interpretation of meaningful classes. To avoid local maxima, the model was estimated with 300 random starting seeds and the maximum number of iterations was set to 100,000 (Lanza et al., 2015).

The decision for the number of classes to retain was based on both the absolute model fit, assessed using G^2^, the likelihood ratio statistic that provides an estimate of how well the data is represented by the model, and the relative model fit, referring to the use of information criterion (Collins & Lanza, 2010), such as the Akaike Information Criterion (AIC: Akaike, 1974), Bayesian Information Criteria (BIC: Schwartz, 1978), adjusted BIC (Sclove, 1987) and consistent AIC (Bozdogan, 1987). Additionally, the entropy diagnostic statistic estimates how accurately the model classifies the latent classes with values closer to 1 indicative of a better model fit (Wang et al., 2017).

Additionally, model selection was based on whether the classes demonstrated adequate homogeneity, referring to the probability that class members would provide homogenous patterns of HRBs, and adequate separation, referring to the degree to which the response patterns differ between classes (Collins & Lanza, 2010). Both homogeneity and separation of the latent classes was determined using the rho parameters, which indicate the likelihood that an individual would endorse a HRB given their class membership. In the current study, rho parameters above .5 were indicative of a high probability that class members would engage in a particular HRB. Finally, the gamma parameters provided an estimate of the size of the latent classes (Collins & Lanza, 2010).

#### Conditional model

After selecting the optimal number of latent classes, the covariates tested independently for significance. Multicollinearity of the predictors was assessed, and highly correlated covariates were excluded from the analysis to protect against family wise error. The continuous covariates were standardised prior to their entry into the model. Covariates with more than two categories were dummy coded. The significant covariates were then added to the multinomial logistic regression model in hierarchical order, starting with the demographic variables, then ACE scores, followed by the remaining covariates.

## Results

### Preliminary Analysis

#### Missing Data

SPSS and the PROC LCA procedure in SAS 9.4 were used to conduct the analysis. Missing data was assumed to be missing at random (MAR) (Centre for Longitudinal Studies, 2010). The MCS has a complex sample design as the sample was geographically clustered and disproportionately stratified to ensure that areas high in child poverty and minority ethnic groups would be overrepresented (Centre for Longitudinal Studies, 2010). Despite recommendations to incorporate these design features and use inverse probability weights to account for non-response, the chosen analytical software does not currently allow for these design features to be considered in the analysis, which may lead to biased parameter estimates.

Full information maximisation likelihood estimation (FIML) was used, allowing for missing data on the HRB indicators to be included in the analysis as FIML utilises all of the data in the sample to produce adjusted parameter estimates (Enders & Bandolos, 2001). However, the FIML procedure cannot analyse missing information on the covariates, resulting in cases with missing data on the covariates being removed from the analysis when covariates are included in the model. Overall, 497 cohort members were missing information on at least one covariate and therefore were excluded from the analysis.

#### Descriptive Statistics

Descriptive statistics for the study variables are presented in **Table 1**. The average age of the sample at T1 was 37.26 (*SD* = 15.29), at T2 the average age was 37.33 (*SD* = 15.21) and at T3 the average age was 37.59 (*SD* = 15.06). The sample was majority female (68%) and white (95%). Around 48% of the sample reported having no academic qualifications.

As shown in Table 2, the most frequent ACE reported was parental substance use (16%) and physical assault (16%) and around 71% of the sample reported experiencing at least 1 ACE. The scores on the UCLA loneliness scale were relatively low at T2 (*M* = 2.13, *SD* = 2.32), which may reflect the lack of lockdown restrictions at this timepoint.

**Table 2.** Prevalence of ACEs.

| Adverse Childhood Experience | BCS70 |  | MCS |  | Full Sample |  |
| --- | --- | --- | --- | --- | --- | --- |
|  | n | % | n | % | n | % |
| Suboptimal Parenting Style | 242 | 14.40 | 218 | 26.20 | 460 | 12.60 |
| Parental Substance Use | 568 | 33.80 | 33 | 2.90 | 601 | 16.40 |
| Incarceration | 262 | 15.60 | 8 | 0.70 | 270 | 7.40 |
| Exposure to Domestic Violence | - |  | 28 | 2.50 | 28 | 0.77 |
| Poor Parental Mental Health | 221 | 13.20 | 215 | 18.50 | 436 | 11.90 |
| Physical Punishment | 33 | 2.00 | 135 | 12.20 | 168 | 4.60 |
| Victimisation (Sexual Assault) | 185 | 11.00 | 37 | 3.20 | 222 | 6.10 |
| Victimisation (Physical Assault) | 83 | 4.90 | 220 | 18.90 | 303 | 8.30 |
| Victimisation (Verbal Assault) | 80 | 4.80 | 512 | 44.10 | 592 | 16.20 |
| Parental Separation | 296 | 17.60 | 177 | 15.90 | 473 | 12.90 |
| Parental Death | 80 | 4.80 | 10 | 0.90 | 90 | 2.50 |
| Foster care | 15 | 0.90 |  |  | 15 | 0.41 |
| ACE Category |  |  |  |  |  |  |
| 0 ACEs | 525 | 31.30 | 312 | 26.70 | 837 | 29.40 |
| 1 ACE | 699 | 41.60 | 398 | 34.10 | 1097 | 38.50 |
| 2 ACEs | 309 | 210.20 | 263 | 135.60 | 572 | 20.10 |
| 3+ ACEs | 147 | 8.80 | 194 | 16.60 | 341 | 12.00 |

*Note.* ACE = Adverse childhood experiences.

**Table 3.** Frequencies and Chi Square Results for HRB Items (N = 2,847)

| Health Behaviour |  | Wave 1 |  |  |  | Wave 2 |  | X<sup>2</sup> |
| --- | --- | --- | --- | --- | --- | --- | --- | --- |
|  |  | n | % | n | % | n | % |  |
| BMI |  |  |  |  |  |  |  |  |
|  | Overweight/Obese BMI | 1091 | 47.89 | 1111 | 48.35 | 1100 | 49.62 | .489 |
|  | Healthy/Underweight BMI | 1187 | 52.11 | 1187 | 51.65 | 1117 | 50.38 |  |
| Smoke Status |  |  |  |  |  |  |  |  |
|  | Current Smoker | 299 | 10.52 | 410 | 14.43 | 330 | 11.91 | 20.70* |
|  | Non/Ex-Smoker | 2543 | 89.48 | 2431 | 85.57 | 2440 | 88.09 |  |
| Sleep Duration |  |  |  |  |  |  |  |  |
|  | Less than 7/More than 9 | 1887 | 66.42 | 1847 | 65.10 | 1785 | 64.63 | 2.145 |
|  | Between 7 and 9 Hours | 954 | 33.58 | 990 | 34.90 | 977 | 35.37 |  |
| Frequency of Alcohol Consumption |  |  |  |  |  |  |  |  |
|  | 4 or more times a week | 661 | 25.90 | 665 | 28.36 | 564 | 23.87 | 12.36* |
|  | Less than 4 times a week | 1891 | 74.10 | 1680 | 71.64 | 1799 | 76.13 |  |
| Average Number of Drinks |  |  |  |  |  |  |  |  |
|  | Binge Drinking Behaviour<sup>b</sup> | 173 | 6.13 | 372 | 13.09 | 300 | 10.83 | 79.163* |
|  | No Binge Drinking | 2650 | 93.87 | 2470 | 86.91 | 2469 | 89.17 |  |

\* $p < .001$ .

### Primary Analysis

#### Unconditional Model

##### Model Identification

Table 4 shows the results from multiple RMLCA with one to nine classes. The frequency distribution of the G^2^ values was examined to assess the percentage of seeds associated with the lowest G^2^ value. The five-class solution had the highest percentage of seeds associated with the smallest G^2^ value, indicating that this value is likely the maximum likelihood solution. Additionally, the frequency distributions of the seven, eight and nine class solutions indicate that the model is under identified.

**Table 4.** Fit Statistics for 1-9 Class Unconditional Models.

| Number<br>of Classes | $G^2$ | $df$ | AIC | BIC | CAIC | ABIC | Entropy | %<sup>a</sup> |
| --- | --- | --- | --- | --- | --- | --- | --- | --- |
| 1 | 14011.86 | 32752 | 14041.86 | 14131.17 | 14146.17 | 14083.51 | 1.00 | 100 |
| 2 | 9602.1 | 32736 | 9664.1 | 9848.68 | 9879.68 | 9750.18 | .87 | 44.50 |
| 3 | 7286.94 | 32720 | 7380.94 | 7660.78 | 7707.78 | 7511.44 | .91 | 37.50 |
| 4 | 5677.42 | 32704 | 5803.42 | 6178.52 | 6241.52 | 5978.35 | .91 | 69.00 |
| **5** | **4729.06** | **32688** | **4887.06** | **5357.43** | **5436.43** | **5106.42** | **.89** | **80.00** |
| 6 | 4265.62 | 32672 | 4455.62 | 5021.26 | 5116.26 | 4719.41 | .87 | 38.50 |
| 7 | 3838.28 | 32656 | 4060.28 | 4721.18 | 4832.18 | 4368.49 | .87 | 16.50 |
| 8 | 3561.03 | 32640 | 3815.03 | 4571.19 | 4698.19 | 4167.67 | .88 | 6.50 |
| 9 | 3296.44 | 32624 | 3582.44 | 4433.86 | 4576.86 | 3979.5 | .85 | 7.50 |

*Note.* Bold indicates chosen model.
<sup>a</sup>Percentage of seeds associated with best fitted model.

##### Model Selection

The model fit statistics of the nine candidate models were compared to find the optimal solution. The item response probabilities for the four, five and six class solutions were examined. When the three models were compared, despite the six-class solution having a lower AIC, BIC, ABIC and CAIC, the five-class solution provided a more parsimonious explanation of the differing patterns and trajectories of response, and the latent classes had a more meaningful interpretation compared to the four-class solution. The entropy value for the five-class solution was adequate (.89) indicating that the latent classes were well classified.

Table 5 reports the marginal proportions, and the parameter estimates for class prevalence and item response probabilities. The overall probability of cohort members reporting a high BMI was consistent across the three timepoints, with a slight increase from T1 to T3 (.48 - .50). Initially, the overall probability of cohort members smoking was low (.11), but increased at T2 (.11 - .14). The elevated probability reduced at T3 but remained higher than at T1 (.14 - .12). There was a consistently high probability of reporting atypical sleep that remained stable across the three timepoints (.66-.65). The probability of reporting drinking more than 4 times a week was relatively low at T1 (.26), but this increased slightly at T2 (.28) before reverting to a lower probability of frequent alcohol consumption at T3 compared to T1 (.24). Finally, the probability of binge drinking was low across the three timepoints but doubled from T1 to T2 (.06 - .13), with this elevated likelihood of binge drinking still present at T3 (.13-.11). The following section described how the latent classes were described and labelled.

**Table 5.**
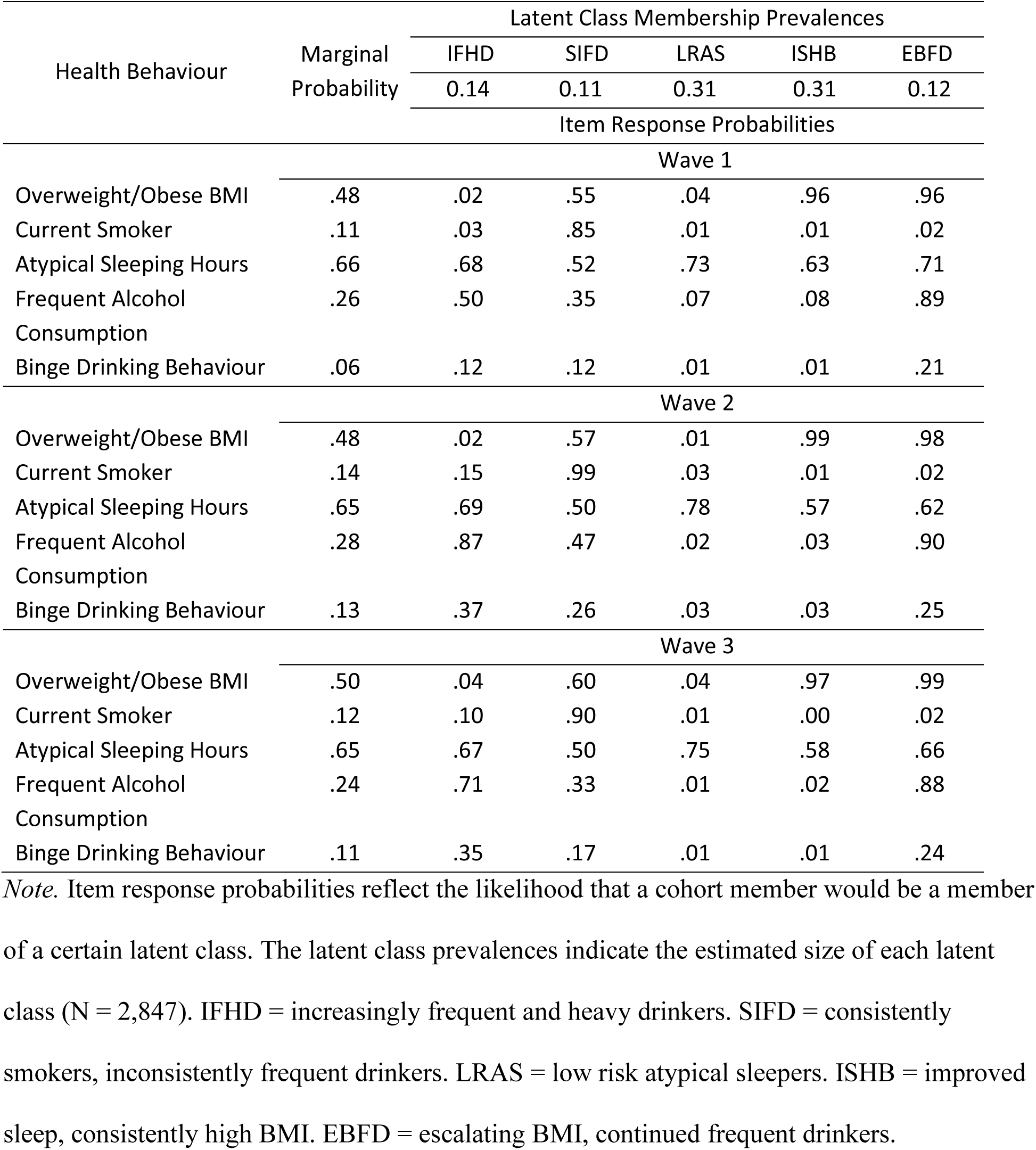
Item Response Table.

#### Description of Latent Classes

##### Increasingly Frequent and Heavy Drinkers

Approximately 14% of the sample (*N* = 411) comprised the “*Increasingly Frequent and Heavy Drinkers”.* Class members had a very low probability of indicating a high BMI across the three timepoints (.02- .04) and there was a high probability of atypical sleep that remained stable across time (.68 - .67). Overall class members had a very low probability of reporting smoking across the three timepoints, however, there was an increased probability from T1 to T2 (.03 - .15) that decreased at T3, but the T3 likelihood of class members smoking remained higher compared to T1 (.10). Class members had an initially high probability of drinking more than 4 times a week (.50), which increased by approximately 37% at T2. While the probability of frequent drinking at T3 decreased from that at T2, there was still a 21% higher likelihood of frequent alcohol consumption at T3 compared to at T1. Class members had a low probability of engaging in binge drinking across the three timepoints (< .5), however, at T2 the likelihood of binge drinking rose by 15% and remained higher at T3 compared to T1.

**Figure 2.**
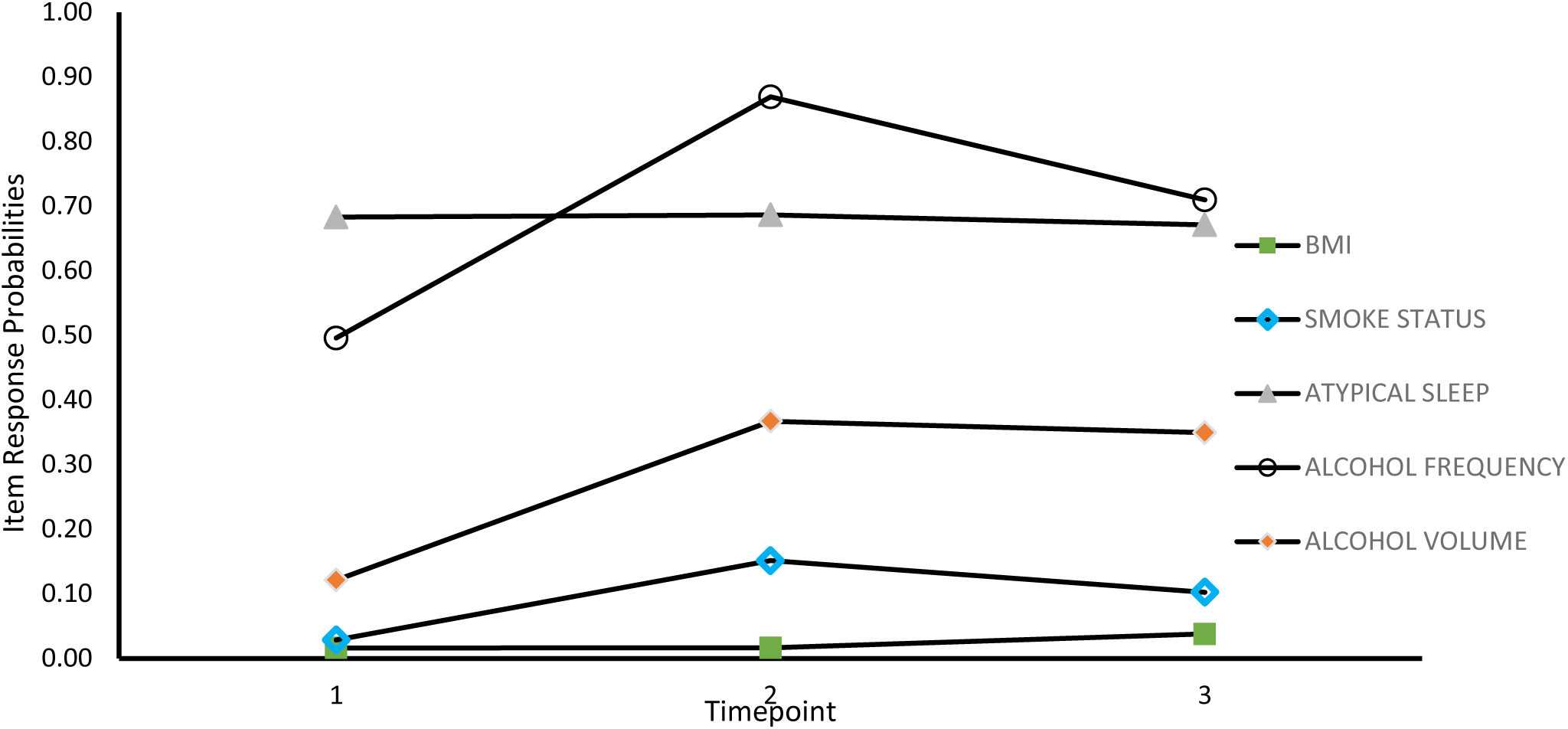
Plot of Item Response Probabilities for IFHD.

##### Consistently Smokers, Inconsistently Frequent Drinkers

Approximately 11% (*N* = 313) comprised the second class labelled *Consistently Smokers, Inconsistently Frequent Drinkers (SIFD)* (Figure 3). The probability of smoking was initially high at T1 (.85) and showed an upwards trajectory from T1 to T2 (.99) before declining to above T1 probabilities at T3 (.90). Class members had relatively stable high probability of reporting atypical sleep that reduced slightly across the three timepoints (.52-.50). The probability of reporting a high BMI was initially high (.55) and showed a slight upwards trajectory from T2 (.57) to T3 (.60). Despite the low probability of drinking more than four times a week at T1 (.35), the likelihood of frequent drinking increased by 12% at T2, before reverting to below the T1 probability (.33). Similarly, the probability of binge drinking at T2 was approximately double that at T1 (.12 - .26) and showed a downwards trajectory from T2 to T3, but the likelihood of binge drinking was still higher at T3 compared to T1 (.17). Overall, class members showed an increased probability of smoking and problematic alcohol consumption across the three timepoints.

**Figure 3.**
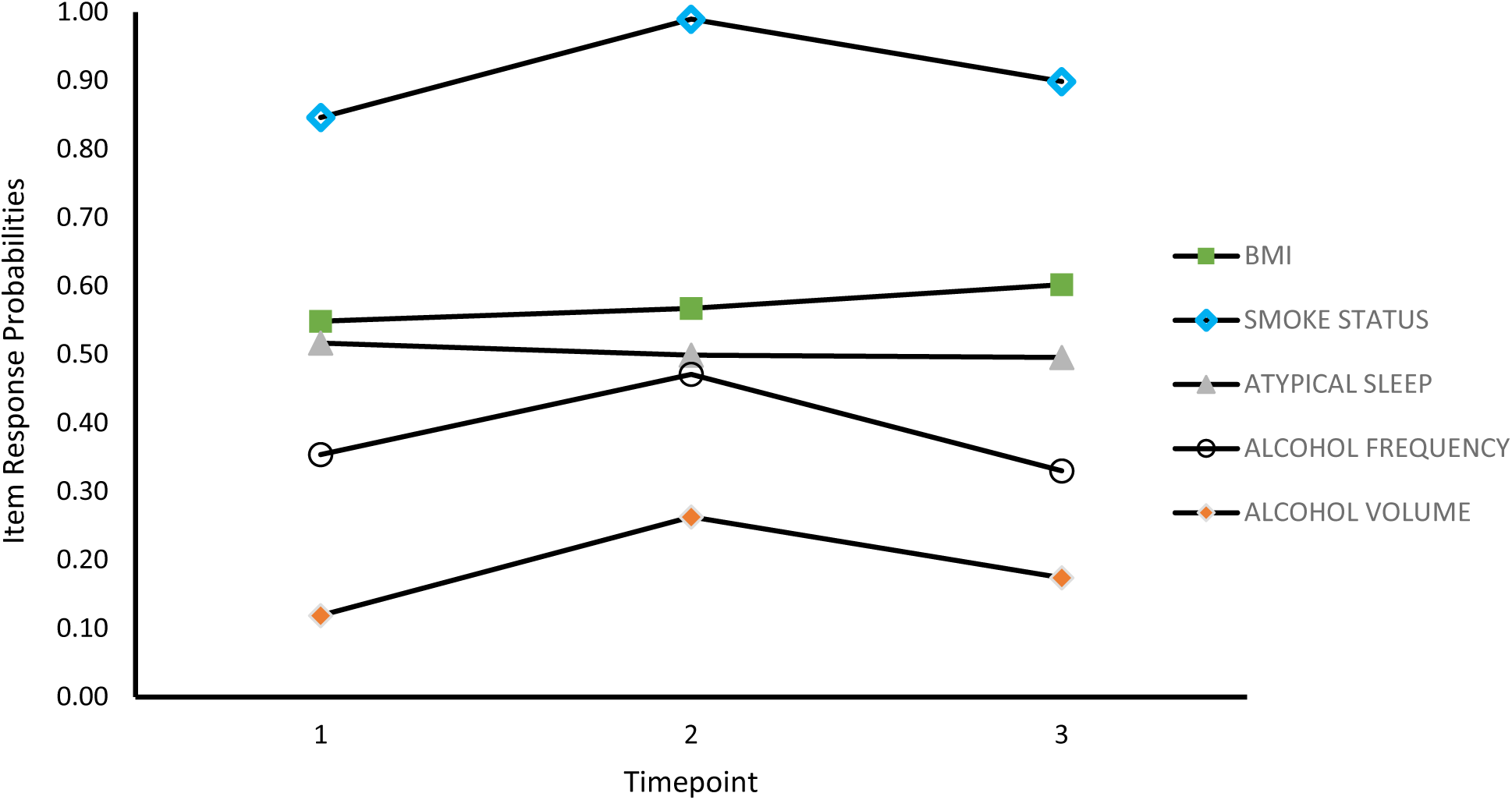
Plot of Item Response Probabilities for SIFD.

##### Low Risk Atypical Sleepers

Approximately 31% (*N* = 893) of the sample were members of the third class labelled *low risk atypical sleepers (LRAS)* (Figure 4). The item response probabilities for cohort members reporting smoking, frequent drinking, binge drinking, or a high BMI was consistently very low across the three timepoints. Class members had a high likelihood of reporting atypical sleep at T1 (.73) which showed a slight upwards trajectory at T2 (.78) before reverting to near T1 levels at T3 (.75).

**Figure 4.**
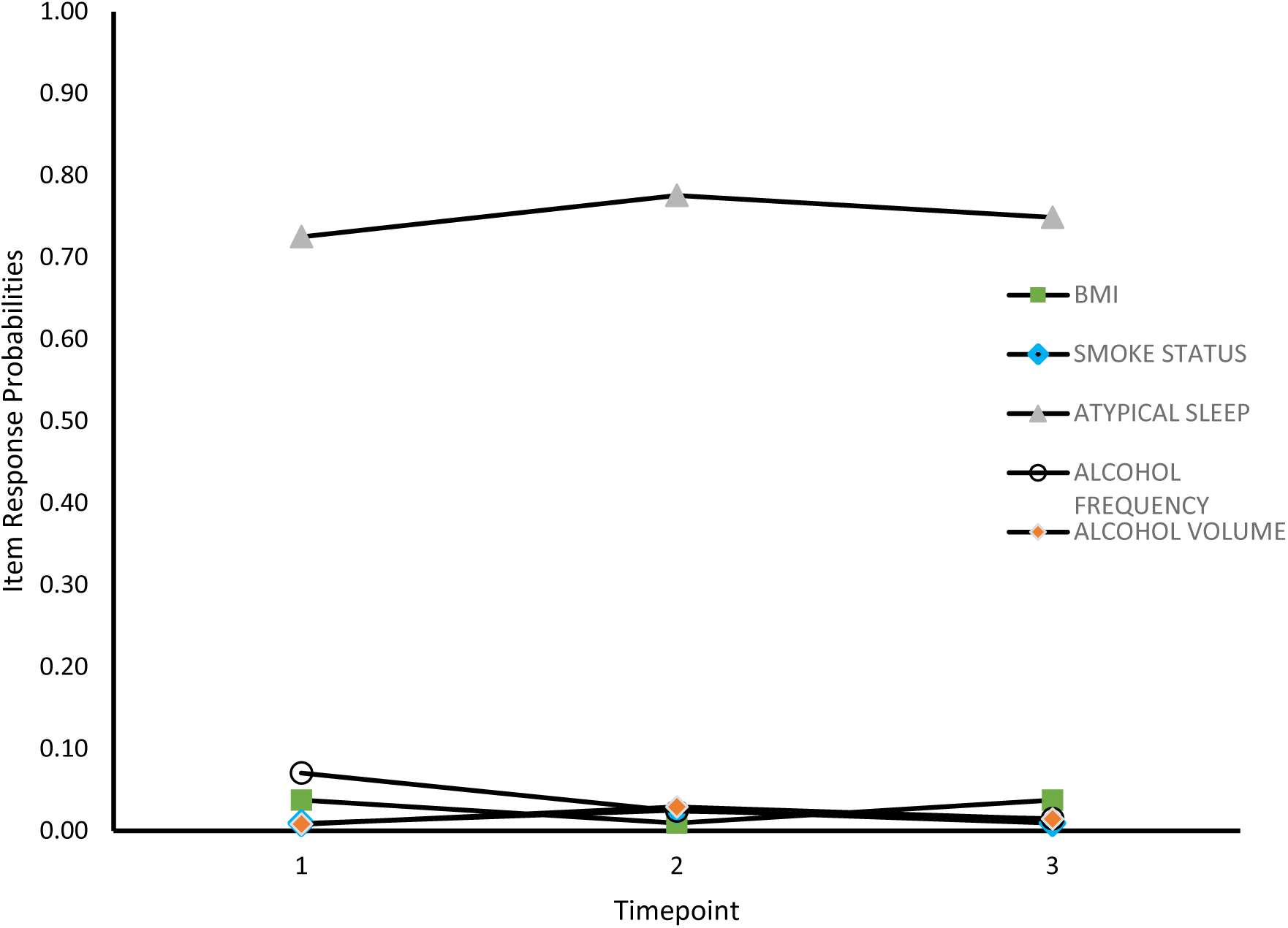
Plot of Item Response Probabilities for LRAS.

##### Improved Sleep, Consistently High BMI

Approximately 31% (*N* = 880) of the sample comprised the fourth class labelled *improved sleep, consistently high BMI* (ISHB) (Figure 5), characterised by a high probability of reporting a high BMI at T1 (.96) that increased slightly at T2 (.99) before reverting to near T1 levels at T3 (.97). Class members has a near 0% likelihood of reporting smoking, that remained stable across the three timepoints. Unlike the other classes, this class demonstrates a downwards trajectory in the probability that they drink more than 4 times a week (.08-.02). The likelihood of binge drinking showed a slight upwards trajectory from T1 to T2 (.01 - .03) before reverting to the probability of binge drinking at T1. Class members showed an initial high likelihood of reporting atypical sleep (.63) which showed a downwards trajectory from T1 to T2 (.57), and the attenuated likelihood of atypical sleep duration remained stable at T3 (.57).

**Figure 5.**
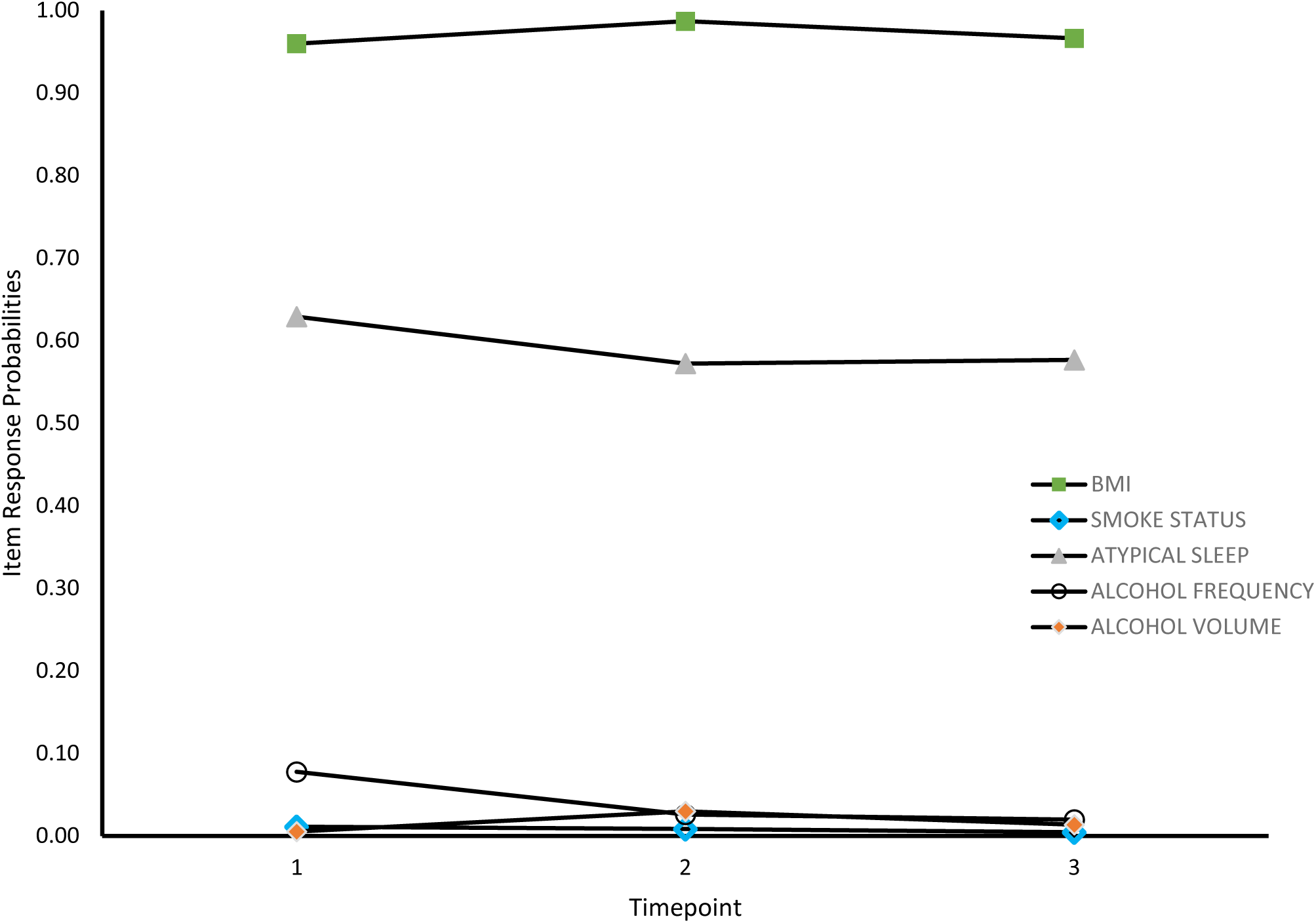
Plot of Item Response Probabilities for ISHB.

##### Escalating BMI, Continued Frequent Drinkers

The last class comprised approximately 12% of the sample (N = 350) and was labelled *Escalating BMI, Continued Frequent Drinkers (EBFD)* (Figure 6). The probability of reporting a high BMI was initially high (.96) and showed an upwards trajectory across the subsequent timepoints, cumulating in a 99% probability of class members reporting a high BMI at T3. The low likelihood of class members smoking remained stable across the three timepoints (.02). Initially, there was a high probability of atypical sleep at T1 (.71) but this showed a downwards trajectory from T1 to T2 (.71 - .62). The probability of atypical sleep increased slightly at T3 but remained below the probability at T1 (.66). As for alcohol consumption, the probability of binge drinking was the highest among the five classes at T1 (.21) but displayed a less drastic upwards trajectory at T2 compared to other classes (.21-.25) which remained elevated from T1 levels at T3 (.24). Finally, the probability of class members drinking more than 4 times a week remained consistently high from T1 to T2 (.89-.90) and from T2 to T3 (.90 - .88).

**Figure 6.**
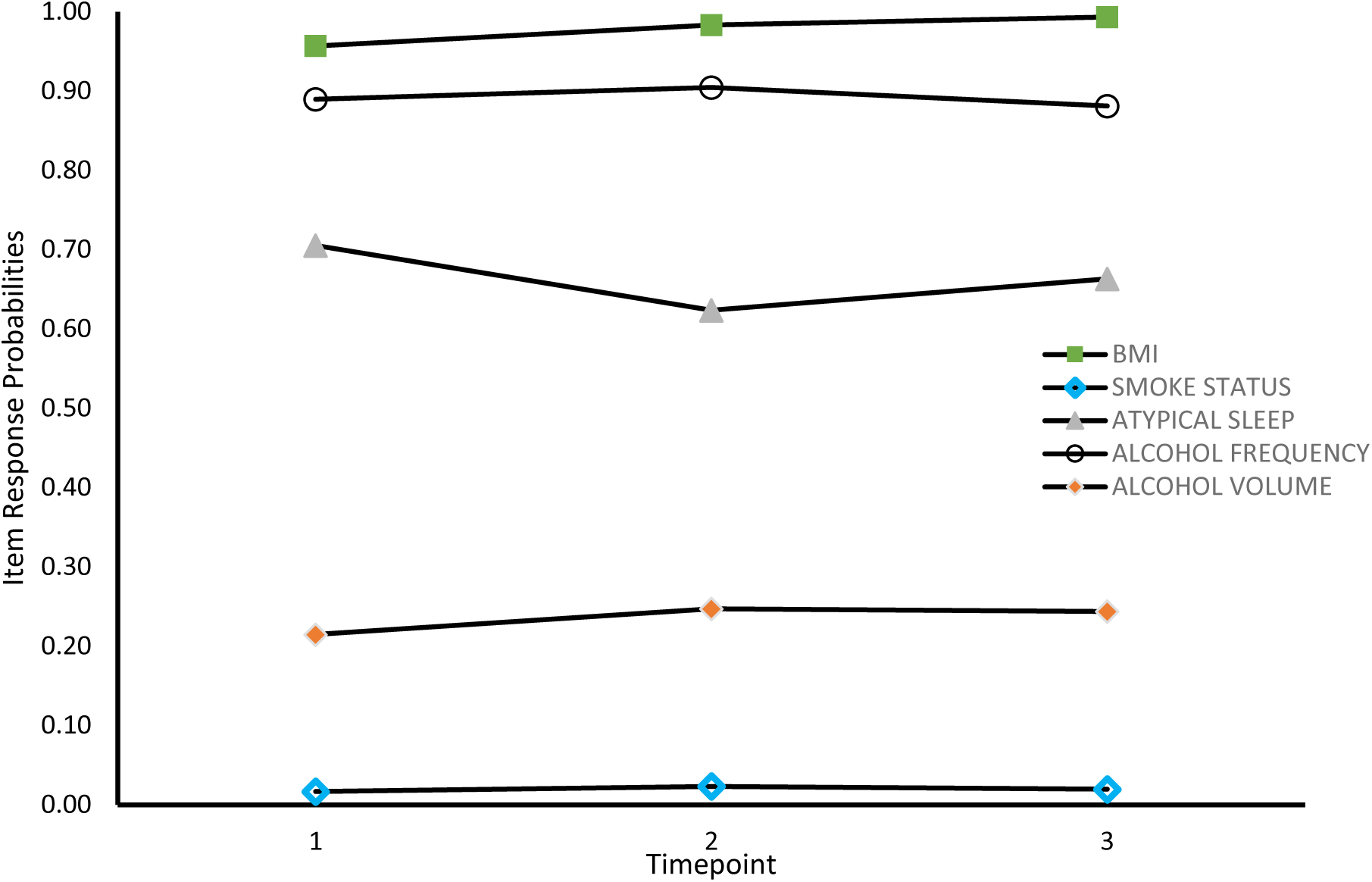
Plot of Item Response Probabilities for EBFD.

#### Conditional Model

Table 6 shows the descriptive statistics and results from a series of chi square tests performed to examine the relationship between the candidate predictors of class membership at each timepoint. Appendix C illustrates the correlation matrix from a series of bivariate correlations that were conducted to assess for multicollinearity between the candidate predictor variables included in the conditional model. Pre-pandemic psychological distress was highly correlated with age (*r* = - .77, *p* < .001), to protect against family wise error, and considering the difference in age between the two cohorts in the sample, pre-pandemic psychological distress was not included as a covariate in the model. Age was subsequently dichotomised to reflect the age groups of the two cohorts. As the scores on the UCLA loneliness scale at the three timepoints were highly correlated with one another (*r* = .72, *p* < .001), only the T2 score was included in the model as this timepoint reflected the highest likelihood of engaging in risky HRBs across multiple classes.

**Table 6.** Frequencies and Chi Square Results for Covariates.

| Health Behaviour | Wave 1 | | Wave 2 | | Wave 3 | | $\chi^2$ |
| --- | --- | --- | --- | --- | --- | --- | --- |
|  | *n* | % | *n* | % | *n* | % |  |
| Instrumental Support |  |  |  |  |  |  |  |
| High | 2476 | 86.97 | 2420 | 85.00 | 2460 | 87.14 | 7.27* |
| Low | 371 | 13.03 | 427 | 15.00 | 363 | 12.86 |  |
| Emotional Support |  |  |  |  |  |  |  |
| High | - |  | 2452 | 86.13 | 2461 | 86.44 | 0.12 |
| Low | - |  | 395 | 13.87 | 386 | 13.56 |  |
| Mental Health Diagnosis |  |  |  |  |  |  |  |
| Yes | 381 | 13.38 | - |  | 534 | 18.76 | 30.48* |
| No | 2466 | 86.62 | - |  | 2313 | 81.24 |  |
| Psychological Distress<sup>a</sup> |  |  |  |  |  |  |  |
| High | 525 | 18.44 | 581 | 20.41 | 589 | 20.69 | 5.37 |
| Low | 2322 | 81.56 | 2266 | 79.59 | 2258 | 79.31 |  |
| Anxiety/Depressive Symptomatology<sup>b</sup> |  |  |  |  |  |  |  |
| No | 2395 | 84.72 | 2388 | 83.94 | 2337 | 82.12 | 7.39* |
| Yes | 432 | 15.28 | 457 | 16.06 | 509 | 17.88 |  |
| Adverse Life Events |  |  |  |  |  |  |  |
| Yes | - |  | 1232 | 43.27 | 1566 | 55.01 | 78.39* |
| No | - |  | 1615 | 56.73 | 1281 | 44.99 |  |
| Social Contact |  |  |  |  |  |  |  |
| High | 2350 | 82.57 | 2233 | 78.43 | 2062 | 74.39 | 55.76* |
| Low | 496 | 17.43 | 614 | 21.57 | 710 | 25.61 |  |

*Note.* Dashes indicate waves where cohort members were not asked the question.
<sup>a</sup> Measured by the Kessler (6) and Malaise Inventory for each cohort respectively.
<sup>b</sup> Measured using the combined 4 item Generalised Anxiety Disorder and Patient Health Questionnaire.

Multinomial logistic regression models were conducted to investigate the relation between covariates and the probability of class membership. The LRAS class was selected as the reference class as class members displayed the lowest likelihood of engaging in risky HRBS. The likelihood ratio test of significance indicated whether each covariate contributed significantly to the prediction of class membership. Due to the large number of covariates, only significant predictors of class membership were reported (Table 7). Appendix D presents the odds ratio and confidence intervals for the full unconditional model.

**Table 7.** Significant Predictors of Class Membership.

|  | IFHD<br>[vs LRAS] |  | SIFD<br>[vs LRAS] |  | ISHB<br>[vs LRAS] |  | EBFD<br>[vs LRAS] |  |
| --- | --- | --- | --- | --- | --- | --- | --- | --- |
|  | OR | (95% CI) | OR | (95% CI) | OR | (95% CI) | OR | (95% CI) |
| Covariates measured pre-pandemic |  |  |  |  |  |  |  |  |
| Age group* |  |  |  |  |  |  |  |  |
| 18-20 | (ref) |  | (ref) |  | (ref) |  | (ref) |  |
| 49-50 | 2.38 | [1.51 , 3.75] | 1.96 | [1.27 , 3.02] | 5.07 | [3.66 , 7.04] | 8.05 | [5.21 , 12.43] |
| Gender* |  |  |  |  |  |  |  |  |
| Male | (ref) |  | (ref) |  | (ref) |  | (ref) |  |
| Female | 0.91 | [0.69 , 1.19] | 0.69 | [0.54 , 0.89] | 0.68 | [0.56 , 0.82] | 0.45 | [0.36 , 0.57] |
| Race* |  |  |  |  |  |  |  |  |
| Minority Group | (ref) |  | (ref) |  | (ref) |  | (ref) |  |
| White | 0.49 | [0.31 , 0.79] | 1.52 | [0.84 , 2.77] | 0.67 | [0.44 , 1.01] | 1.62 | [0.76 , 3.43] |
| ACE Category |  |  |  |  |  |  |  |  |
| 0 ACEs | (ref) |  | (ref) |  | (ref) |  | (ref) |  |
| 2 ACEs | 1.18 | [0.84 , 1.67] | 1.77 | [1.29 , 2.41] | 1.08 | [0.85 , 1.38] | 1.04 | [0.76 , 1.43] |
| Covariates measured during T1 |  |  |  |  |  |  |  |  |
| Psychological distress |  |  |  |  |  |  |  |  |
| No | (ref) |  | (ref) |  | (ref) |  | (ref) |  |
| Yes | 1.25 | [0.84 , 1.85] | 1.57 | [1.07 , 2.29] | 1.67 | [1.22 , 2.29] | 2.47 | [1.68 , 3.64] |
| Covariates measured during T2 |  |  |  |  |  |  |  |  |
| Adverse life events |  |  |  |  |  |  |  |  |
| No | (ref) |  | (ref) |  | (ref) |  | (ref) |  |
| Yes | 1.28 | [0.99 , 1.66] | 1.43 | [1.12 , 1.83] | 0.96 | [0.79 , 1.16] | 1.24 | [0.98 , 1.58] |
| Loneliness score | 1.27 | [1.09 , 1.48] | 1.09 | [0.94 , 1.26] | 1.20 | [1.07 , 1.34] | 0.91 | [0.78 , 1.06] |
| Covariates measured during T3 |  |  |  |  |  |  |  |  |
| Adverse life events |  |  |  |  |  |  |  |  |
| No | (ref) |  | (ref) |  | (ref) |  | (ref) |  |
| Yes | 0.93 | [0.72 , 1.21] | 1.13 | [0.88 , 1.44] | 1.37 | [1.14 , 1.65] | 1.25 | [0.98 , 1.59] |

*Note.* LRAS represents the reference class. ACE = Adverse childhood experiences. \* $p < .0001$ .

##### Baseline Covariates

As shown in Table 7, the model showed that age significantly predicted class membership and the odds ratios indicate that those ages 49-50 were 2.4 times more likely to be members of the IFHD class, almost twice as likely to be in the SIFD class, 5.1 times more likely to be in the ISHB class and 8.1 times more likely to be in the EBFB class. Suggesting that older cohort members were more likely to be members of a class that had a high probability of endorsing at risk health behaviours.

Gender was also a significant predictor of class membership, and the odds ratios show that females were 9% more likely to be members of the LRAS class than the IFHD class, 31% more likely to be in the LRAS class compared to the SIFD class, 32% more likely to be members of the LRAS compared to the ISHB class and 55% more likely to be members of the LRAS class compared to the EBFD class. Suggesting that females were more likely to be members of the latent class with the lowest probability of engaging in risky HRBs.

Race was a significant predictor of class membership, with white cohort members 51% more likely to be in the LRAS class compared to the IFHD class, 52% more likely to be members of the SIFD class compared to the LRAS class, 33% more likely to be members of the LRAS class compared to the ISHB class and 62% more likely to be in the EBFD class compared to the LRAS class. Suggesting that white cohort members were more likely to be a member of a latent class that more than one risky HRB.

A history of ACEs was only significant for those with an ACE score of 2, *p* = .048. Experiencing two ACEs was predictive of a 18% higher likelihood of membership in the IFHD class, 77% higher likelihood of being a member of the SIFD class, 8% higher for membership in the ISHB latent class and 4% higher probability for membership in the EBFD latent class compared to the LRAS class, suggesting that a history of two ACES is the strongest predictor for membership in the SIFD class who have the highest probability of smoking.

##### T1 Covariates

Psychological distress at T1 was significantly predictive of class membership *p* = .0015, with those reporting significant levels of psychological distress 25% more likely to be members of the IFHD class, 57% more likely to be members of the SIFD class, 67% more likely to be members of the ISHB class and 2.47 times more likely to be members of the EBFD class compared to the LRAS class.

##### T2 Covariates

At T2 both loneliness, *p* = .0087 and adverse life experiences during the pandemic, *p* = 0.0361 were predictive of class membership. Experiencing adverse life events was predictive of a 28% higher likelihood of being a member of the IFHD class, 43% higher probability of membership in the SIFD class, a 24% higher likelihood of membership in the EBFD class and a 9% lower probability of membership in the ISHB class compared to the LRAS class. Suggesting that adverse life events increased the likelihood that cohort members would be members of a latent class that endorsed risky HRBs but decreased the likelihood they would be members of the ISHB class.

For every unit increase on the UCLA loneliness scale, the probability of membership in the IFHD class increased by 27% compared to the LRAS class. There was a 9% higher likelihood of membership in the SIFD class, 20% higher likelihood of membership in the ISHB class but a 9% reduced likelihood of membership in the EBFD class compared to the LRAS class. Suggesting that high loneliness at T2 was predictive of a higher probability of engaging in risky HRBs but a decreased likelihood of reporting both a high BMI and frequent alcohol use.

##### T3 Covariates

Finally, at T3 adverse life events was predictive of 13% higher probability of membership in the SIFD class, 37% higher likelihood of membership in the ISHB class and 25% higher probability of membership in the EBFD class compared to the LRAS class. However, the probability of membership in the IFHD compared to the LRAS class reduced by 7% suggesting that adverse life events at T3 was less predictive of membership in the IFHD class compared the 27% increased probability of membership at T2.

## Discussion

The current study examined the trajectory of patterns of HRBs during the first nine months of the covid-19 pandemic. We identified five distinct subgroups of individuals with similar patterns of HRBs and demonstrated how the adoption of risky HRBs differed between the classes. While we were unable to assess whether individuals transitioned between subgroups across time and the factors that predicted this transition, we were able to use longitudinal data to ascertain how differing groups changed their HRBs over time and examine the psychosocial factors that were predictive of group membership. We hypothesised that various risk and resilience factors would predict class membership, however, in contrast to previous studies, our findings did not show any relationship between risky HRBs, social support and poor mental health during the pandemic (Villadsen, 2020).

### Summary of Findings

The current study found that a history of two ACEs was predictive of membership in classes with a higher probability of risky health behaviours. The relationship between ACEs and a higher likelihood of engaging in maladaptive HRBs has been illustrated in previous studies (Wiss & Brewerton, 2020), however, unlike with those findings, we did not find evidence to support a cumulative effect of ACEs on an increased susceptibility to risky HRBs. For instance, while we did find an increased probability of membership in the EBFD class reflecting findings from previous research indicating a 46% higher likelihood of obesity amongst those with ACEs, their results showed a dose-dependent relationship between weight gain and ACE exposure that was not replicated in the current study (Mahmouud et al., 2022). A possible explanation for this is the use of single items to measure ACEs. In fact, as shown in Appendix B, many indicators of exposure were measured at different ages in the two cohorts, which may have implications regarding the impact of ACEs on later health outcomes due to the differential effect of ACEs depending on the age at which they were experienced (Atzl et al., 2019).

Atypical sleep duration was a consistent finding across all classes, and the detrimental impact of the pandemic on sleep is supported by finding from international studies that found an increase in sleep disturbances worldwide during the pandemic (Jahrami et al., 2022). Restrictions on movement, reduced contact with loved ones and a lack of positive stimulation in their environment meant that people were more likely to spend more time on their phones (Altena et al., 2020). Thus, as illustrated by Sultana et al., (2021) who found an association between reported screentime and poor sleep quality during the pandemic, increased screentime may be a causal factor explaining the current study’s finding of sleep disturbances across all classes.

A prior study used latent transition analyses to investigate the adoption of risky HRBs over a similar time period (July 2020 and November 2020) during the pandemic in Chile (Salazar-Fernandez et al., 2022). As with the current study, they were able to identify subgroups of individuals with differing clusters of HRBs, however, unlike with the current study, their findings reflect that the majority of participants either displayed no change in HRBs or transitions to low-risk latent classes. In contrast, while we cannot examine transitions between subgroups in the present study, findings suggest that there was a general increase in the adoption of unhealthy behaviours across time in our sample.

Indeed, the observation that alcohol consumption in the UK took a different course during lockdown compared to other countries has been highlighted in other studies. One large scale study that examined changes in alcohol consumption in 21 European countries found that the UK was the only country where the mean alcohol consumption increased in the first few months of the pandemic (Killian et al., 2021), suggesting that even with the closure of pubs and shops during lockdown, individuals in the UK increased their alcohol consumption.

The findings suggest that the initial adoption of unhealthy behaviours in the current study may have been driven by the motivation to alleviate psychological distress. In fact, psychological distress at T1 significantly predicted a higher likelihood of membership in a latent class likely to adopt unhealthy behaviours, especially for the EBFD class. These results support the assumptions made by the self-medication hypothesis (Khantzian, 2003) as both increased alcohol consumption and emotional eating have been linked with high levels of psychological distress in previous studies. For example, McAtamney et al. (2021) explored the relationship between emotional eating and negative affect during the pandemic in the UK and found an association between depressive symptomatology and emotional eating. In support of this, Dicken et al., (2021) investigated BMI and alcohol consumption during similar time periods as T1 and T2. They found that increases in BMI was associated with increased alcohol consumption, and combined with the finding of the current study that psychological distress was a stronger predictor of membership in the EBHD class relative to the other classes, the results may support the self-medication hypotheses’ assumption that certain individuals engaged in emotional eating and increased alcohol consumption to manage the elevated psychological distress they were experiencing during the first months of lockdown.

Adverse life events at T2, such as unemployment, financial difficulties and the death of loved ones significantly predicted membership in the latent classes more likely to engage in risky HRBs. However, interestingly, psychological distress was not a significant predictor of class membership at T2 or T3, suggesting that perhaps the self-medication hypothesis fails to provide an adequate explanation for the increase in risky health behaviours during T2.

The findings indicate that the change in HRBs was non-linear, as the highest probability of cohort members from several of the classes adopting risky health behaviours was at T2, the only timepoint that was not conducted during lockdown restrictions. At T2 members of the IFHD, SIFD and EBFD classes had a substantially higher probability of drinking more frequently and consuming more alcohol. The context surrounding the time period between T1 and T2 may provide a partial explanation for this finding. From July to September, lockdown restrictions were eased considerably as shops and restaurants opened, however, meeting inside one’s household was still forbidden (Institute for Government Analysis, 2021). Then in August, the government introduced the “eat out to help out” scheme, intended to stimulate the economy by providing a 50% discount on food and non-alcohol drinks at restaurants (HM, 2020).

Despite previous findings indicating that adverse life events would be associated with increased adoption of risky HRBs through eliciting psychological distress (Bell et al., 2021), when the findings are placed in the context surrounding T2, the increases in alcohol consumption and BMI observed may be due to the increased availability as restrictions were eased, allowing people to seek support from friends to alleviate psychological distress. Indeed, social support has been found to help mitigate negative affect during the pandemic (Szkody et al., 2021), and pubs and restaurants provided a place to see loved ones with the restrictions still in place. The non-significant finding for psychological distress at T2, but an increase in alcohol consumption, therefore, could reflect people using social support as a coping strategy and meeting friends in pubs for a drink, rather than attempting to modulate negative affect in isolation.

Research supports the proposition that in the UK, drinking with friends is a common method for obtaining social support. One study that compared Spanish to British drinking habits during the first month of lockdown showed that Britons were much more likely to engage in “virtual pub sessions”, an activity that allowed for people to continue pub traditions at home and receive support from their friends (Rodrigues et al., 2022). The adoption of virtual drinking sessions illustrates how integral drinking has become to the British social life, so when given the opportunity to return to the pub during July and August, many may have received the social support they needed to cope with stress from their friends in-person, an observation supported by findings that virtual contact does not provide as much perceived emotional or instrumental social support compared to in person meetings (Trepte et al., 2015).

People may have found the reduced prices for food especially inviting if they had suffered recent financial hardship, resulting in the consumption of restaurant compared to home cooked food, which research suggests are higher in caloric, fat, salt and sugar content, contributing to weight gain (Goffe et al., 2017). Therefore, the opening of social venues but the continued restrictions on meeting with friends, may have contributed to the increase in alcohol consumption, with the eat out to help out scheme making less healthy meal options more affordable (Brookes, 2021).

### Limitations

Additionally, the elimination of cases with missing data due to non-response may have further biased the results as nonresponse in both cohorts is more likely amongst males from lower socioeconomic backgrounds with younger, less educated parents (Centre for Longitudinal Studies, 2010; Mostafa & Wiggins, 2014). Considering that research suggests that these factors are associated with an increased likelihood of experiencing more ACEs (Marryat & Frank, 2019), the inability to implement clustering and account for non-response may have limited the diversity of the sample.

### Future Directions and Conclusions

To conclude, the current study furthers our understanding of how differing subgroups of people changed their HRBs throughout the course of the pandemic and illustrates the importance of using person-centred methods to investigate behavioural change. Through identifying subgroups with a higher likelihood of engaging in risky HRBs, the findings may facilitate the development of interventions targeted towards individuals likely to be members of latent classes with risky HRBs and inform comprehensive treatments that are tailored to address multiple co-occurring HRBs. Future research should use a more representative sample with validated measures of ACEs, such as the Adverse Childhood Experience Questionnaire (Zarse et al., 2019), to examine whether ACE significantly impacted the adoption of risky health behaviours during lockdown. Additionally, as the current study did not include baseline measures of HRBs, future research should investigate whether pre-pandemic HRBs are predictive of membership in a particular latent class. Furthermore, as the statistical software used in the current study could not accommodate for the use of multiple imputation to replace missing values, future research using more advanced software would facilitate the inclusion of more participants from underrepresented groups in the two cohorts examined.

## Data Availability

All data produced in the present study are available upon reasonable request to the author.

## Appendix A Hypothesis Test of Measurement Invariance for Latent Transition Analysis

**Table 1.** Results from Hypothesis Testing of Measurement Invariance in Latent Transition Analysis.

| Solution | Model | G<sup>2a</sup> | AIC<sup>b</sup> | BIC<sup>c</sup> | df<sup>d</sup> | DiffG<sup>2e</sup> | Diff<sub>df</sub><sup>f</sup> |
| --- | --- | --- | --- | --- | --- | --- | --- |
| 2 Class | 1 | 10646.92 | 10676.92 | 10766.23 | 32752 | 145.32 | 20 |
|  | 2 | 10501.60 | 10571.60 | 10779.99 | 32732 |  |  |
| 3 Class | 1 | 7533.91 | 7591.91 | 7764.57 | 32738 | 195.45 | 30 |
|  | 2 | 7338.46 | 7456.46 | 7807.75 | 32708 |  |  |
| 4 Class | 1 | 6241.33 | 6335.33 | 6615.17 | 32720 | 226.14 | 40 |
|  | 2 | 6015.19 | 6189.19 | 6707.19 | 32680 |  |  |
| 5 Class | 1 | 4604.48 | 4742.48 | 5153.30 | 32698 | 194.87 | 50 |
|  | 2 | 4409.61 | 4647.61 | 5356.14 | 32648 |  |  |

*Note.* Measurement invariance was imposed in model 1 as parameters were restricted to be equal across time. Measurement invariance was not imposed for model 2.
<sup>a</sup>G2 Likelihood ratio difference test statistic.
<sup>b</sup>AIC Akaike Information Criteria.
<sup>c</sup>BIC Bayesian Information Criterion.
<sup>d</sup>df Degrees of freedom.
<sup>e</sup>DiffG<sup>2</sup> Difference in G<sup>2</sup> between model 1 and model 2.
<sup>f</sup>Diff<sub>df</sub> Difference in df between model 1 and model 2.

We initially planned to use a latent transition analysis (LTA) to identify distinct subgroups of individuals with similar HRBs at each timepoint, and if possible, examine the factors that characterise an individual’s transition between latent statuses across time. A hypothesis test of measurement invariance was conducted by comparing models where the rho parameters were freely estimated, to another model where rho parameters were constrained to be equal across time (Collins & Lanza, 2010).

As shown in Table 2, the G^2^ difference test was significant for each of the models tested, indicating that imposing measurement invariance resulted in a significantly weaker model, and the hypothesis of measurement invariance was therefore rejected. As none of the candidate models supported the hypothesis of measurement invariance, this indicates that the meaning of the latent statuses varies across the three timepoints, obscuring the interpretation and labelling of the latent statuses across time. Additionally, as the probability of an individual transitioning across classes over time is conditional on their class membership at the preceding timepoint, the measurement invariance makes characterising the change in status more challenging. Thus, a repeated measures latent class analysis (RMLCA) was ultimately deemed the most suitable analysis as this allowed for the examination of time-dependent patterns of HRBs, without the use of parameter restrictions to impose measurement invariance across time, as is the case with autoregressive models like latent transition analyses.

There are key differences between RMLCA and LTA that should be noted. While both facilitate the identification of distinct subgroups using longitudinal data, rather than characterising how individuals transition between classes across time, RMLCA identifies patterns or trajectories of change across time, and does not provide any estimation of the transition probabilities that would indicate how likely individuals are to transition between classes across time (Lanza & Collins, 2006).

## Appendix B Description of Adverse Childhood Experiences Collected from Both Cohorts

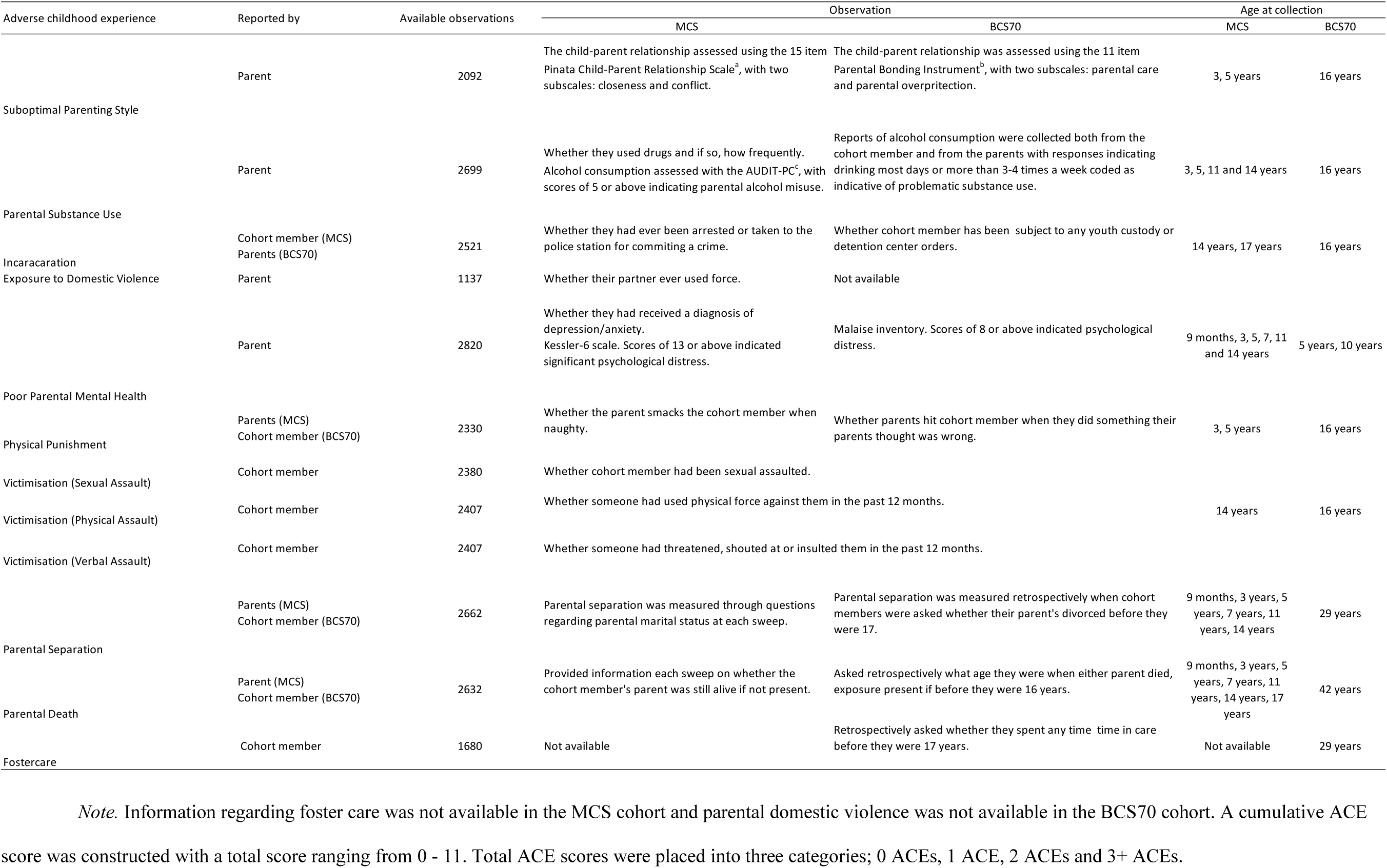
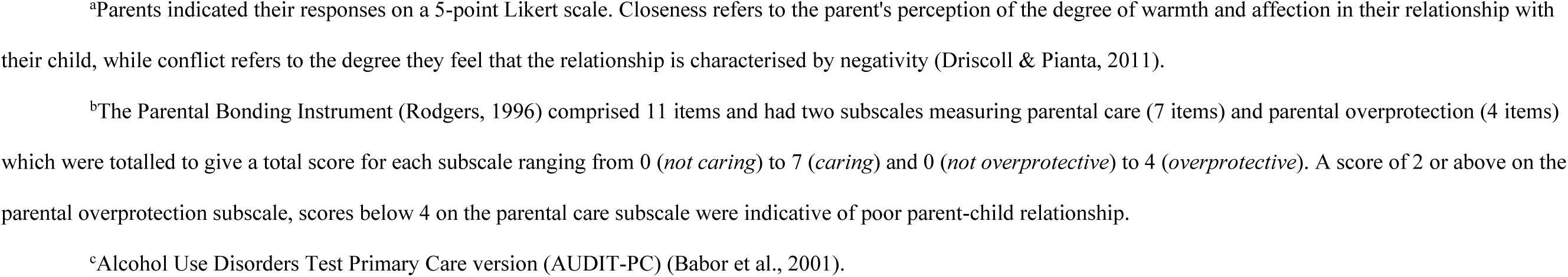

## Appendix C Correlation Matrix of Candidate Covariates

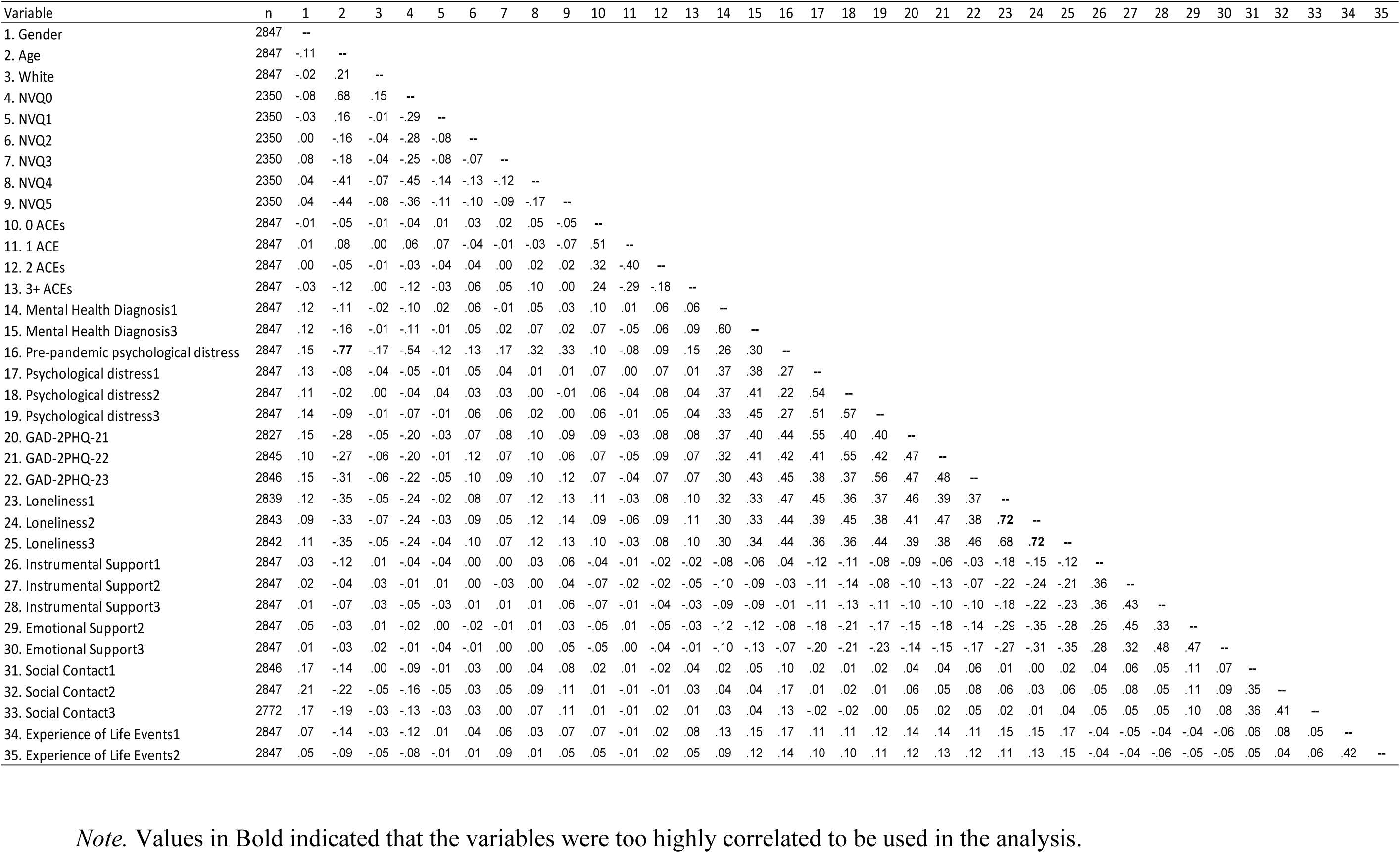

| Variable | n | 1 | 2 | 3 | 4 | 5 | 6 | 7 | 8 | 9 | 10 | 11 | 12 | 13 | 14 | 15 | 16 | 17 | 18 | 19 | 20 | 21 | 22 | 23 | 24 | 25 | 26 | 27 | 28 | 29 | 30 | 31 | 32 | 33 | 34 | 35 |
| --- | --- | --- | --- | --- | --- | --- | --- | --- | --- | --- | --- | --- | --- | --- | --- | --- | --- | --- | --- | --- | --- | --- | --- | --- | --- | --- | --- | --- | --- | --- | --- | --- | --- | --- | --- | --- |
| 1. Gender | 2847 | -- |  |  |  |  |  |  |  |  |  |  |  |  |  |  |  |  |  |  |  |  |  |  |  |  |  |  |  |  |  |  |  |  |  |  |
| 2. Age | 2847 | -.11 | -- |  |  |  |  |  |  |  |  |  |  |  |  |  |  |  |  |  |  |  |  |  |  |  |  |  |  |  |  |  |  |  |  |  |
| 3. White | 2847 | -.02 | .21 | -- |  |  |  |  |  |  |  |  |  |  |  |  |  |  |  |  |  |  |  |  |  |  |  |  |  |  |  |  |  |  |  |  |
| 4. NVQ0 | 2350 | -.08 | .68 | .15 | -- |  |  |  |  |  |  |  |  |  |  |  |  |  |  |  |  |  |  |  |  |  |  |  |  |  |  |  |  |  |  |  |
| 5. NVQ1 | 2350 | -.03 | .16 | -.01 | -.29 | -- |  |  |  |  |  |  |  |  |  |  |  |  |  |  |  |  |  |  |  |  |  |  |  |  |  |  |  |  |  |  |
| 6. NVQ2 | 2350 | .00 | -.16 | -.04 | -.28 | -.08 | -- |  |  |  |  |  |  |  |  |  |  |  |  |  |  |  |  |  |  |  |  |  |  |  |  |  |  |  |  |  |
| 7. NVQ3 | 2350 | .08 | -.18 | -.04 | -.25 | -.08 | -.07 | -- |  |  |  |  |  |  |  |  |  |  |  |  |  |  |  |  |  |  |  |  |  |  |  |  |  |  |  |  |
| 8. NVQ4 | 2350 | .04 | -.41 | -.07 | -.45 | -.14 | -.13 | -.12 | -- |  |  |  |  |  |  |  |  |  |  |  |  |  |  |  |  |  |  |  |  |  |  |  |  |  |  |  |
| 9. NVQ5 | 2350 | .04 | -.44 | -.08 | -.36 | -.11 | -.10 | -.09 | -.17 | -- |  |  |  |  |  |  |  |  |  |  |  |  |  |  |  |  |  |  |  |  |  |  |  |  |  |  |
| 10. 0 ACEs | 2847 | -.01 | -.05 | -.01 | -.04 | .01 | .03 | .02 | .05 | -.05 | -- |  |  |  |  |  |  |  |  |  |  |  |  |  |  |  |  |  |  |  |  |  |  |  |  |  |
| 11. 1 ACE | 2847 | .01 | .08 | .00 | .06 | .07 | -.04 | -.01 | -.03 | -.07 | .51 | -- |  |  |  |  |  |  |  |  |  |  |  |  |  |  |  |  |  |  |  |  |  |  |  |  |
| 12. 2 ACEs | 2847 | .00 | -.05 | -.01 | -.03 | -.04 | .04 | .00 | .02 | .02 | .32 | -.40 | -- |  |  |  |  |  |  |  |  |  |  |  |  |  |  |  |  |  |  |  |  |  |  |  |
| 13. 3+ ACEs | 2847 | -.03 | -.12 | .00 | -.12 | -.03 | .06 | .05 | .10 | .00 | .24 | -.29 | -.18 | -- |  |  |  |  |  |  |  |  |  |  |  |  |  |  |  |  |  |  |  |  |  |  |
| 14. Mental Health Diagnosis1 | 2847 | .12 | -.11 | -.02 | -.10 | .02 | .06 | -.01 | .05 | .03 | .10 | .01 | .06 | .06 | -- |  |  |  |  |  |  |  |  |  |  |  |  |  |  |  |  |  |  |  |  |  |
| 15. Mental Health Diagnosis3 | 2847 | .12 | -.16 | -.01 | -.11 | -.01 | .05 | .02 | .07 | .02 | .07 | -.05 | .06 | .09 | .60 | -- |  |  |  |  |  |  |  |  |  |  |  |  |  |  |  |  |  |  |  |  |
| 16. Pre-pandemic psychological distress | 2847 | .15 | **-.77** | -.17 | -.54 | -.12 | .13 | .17 | .32 | .33 | .10 | -.08 | .09 | .15 | .26 | .30 | -- |  |  |  |  |  |  |  |  |  |  |  |  |  |  |  |  |  |  |  |
| 17. Psychological distress1 | 2847 | .13 | -.08 | -.04 | -.05 | -.01 | .05 | .04 | .01 | .01 | .07 | .00 | .07 | .01 | .37 | .38 | .27 | -- |  |  |  |  |  |  |  |  |  |  |  |  |  |  |  |  |  |  |
| 18. Psychological distress2 | 2847 | .11 | -.02 | .00 | -.04 | .04 | .03 | .03 | .00 | -.01 | .06 | -.04 | .08 | .04 | .37 | .41 | .22 | .54 | -- |  |  |  |  |  |  |  |  |  |  |  |  |  |  |  |  |  |
| 19. Psychological distress3 | 2847 | .14 | -.09 | -.01 | -.07 | -.01 | .06 | .06 | .02 | .00 | .06 | -.01 | .05 | .04 | .33 | .45 | .27 | .51 | .57 | -- |  |  |  |  |  |  |  |  |  |  |  |  |  |  |  |  |
| 20. GAD-2PHQ-21 | 2827 | .15 | -.28 | -.05 | -.20 | -.03 | .07 | .08 | .10 | .09 | .09 | -.03 | .08 | .08 | .37 | .40 | .44 | .55 | .40 | .40 | -- |  |  |  |  |  |  |  |  |  |  |  |  |  |  |  |
| 21. GAD-2PHQ-22 | 2845 | .10 | -.27 | -.06 | -.20 | -.01 | .12 | .07 | .10 | .06 | .07 | -.05 | .09 | .07 | .32 | .41 | .42 | .41 | .55 | .42 | .47 | -- |  |  |  |  |  |  |  |  |  |  |  |  |  |  |
| 22. GAD-2PHQ-23 | 2846 | .15 | -.31 | -.06 | -.22 | -.05 | .10 | .09 | .10 | .12 | .07 | -.04 | .07 | .07 | .30 | .43 | .45 | .38 | .37 | .56 | .47 | .48 | -- |  |  |  |  |  |  |  |  |  |  |  |  |  |
| 23. Loneliness1 | 2839 | .12 | -.35 | -.05 | -.24 | -.02 | .08 | .07 | .12 | .13 | .11 | -.03 | .08 | .10 | .32 | .33 | .47 | .45 | .36 | .37 | .46 | .39 | .37 | -- |  |  |  |  |  |  |  |  |  |  |  |  |
| 24. Loneliness2 | 2843 | .09 | -.33 | -.07 | -.24 | -.03 | .09 | .05 | .12 | .14 | .09 | -.06 | .09 | .11 | .30 | .33 | .44 | .39 | .45 | .38 | .41 | .47 | .38 | **.72** | -- |  |  |  |  |  |  |  |  |  |  |  |
| 25. Loneliness3 | 2842 | .11 | -.35 | -.05 | -.24 | -.04 | .10 | .07 | .12 | .13 | .10 | -.03 | .08 | .10 | .30 | .34 | .44 | .36 | .36 | .44 | .39 | .38 | .46 | .68 | **.72** | -- |  |  |  |  |  |  |  |  |  |  |
| 26. Instrumental Support1 | 2847 | .03 | -.12 | .01 | -.04 | -.04 | .00 | .00 | .03 | .06 | -.04 | -.01 | -.02 | -.02 | -.08 | -.06 | .04 | -.12 | -.11 | -.08 | -.09 | -.06 | -.03 | -.18 | -.15 | -.12 | -- |  |  |  |  |  |  |  |  |  |
| 27. Instrumental Support2 | 2847 | .02 | -.04 | .03 | -.01 | .01 | .00 | -.03 | .00 | .04 | -.07 | -.02 | -.02 | -.05 | -.10 | -.09 | -.03 | -.11 | -.14 | -.08 | -.10 | -.13 | -.07 | -.22 | -.24 | -.21 | .36 | -- |  |  |  |  |  |  |  |  |
| 28. Instrumental Support3 | 2847 | .01 | -.07 | .03 | -.05 | -.03 | .01 | .01 | .01 | .06 | -.07 | -.01 | -.04 | -.03 | -.09 | -.09 | -.01 | -.11 | -.13 | -.11 | -.10 | -.10 | -.10 | -.18 | -.22 | -.23 | .36 | .43 | -- |  |  |  |  |  |  |  |
| 29. Emotional Support2 | 2847 | .05 | -.03 | .01 | -.02 | .00 | -.02 | -.01 | .01 | .03 | -.05 | .01 | -.05 | -.03 | -.12 | -.12 | -.08 | -.18 | -.21 | -.17 | -.15 | -.18 | -.14 | -.29 | -.35 | -.28 | .25 | .45 | .33 | -- |  |  |  |  |  |  |
| 30. Emotional Support3 | 2847 | .01 | -.03 | .02 | -.01 | -.04 | -.01 | .00 | .00 | .05 | -.05 | .00 | -.04 | -.01 | -.10 | -.13 | -.07 | -.20 | -.21 | -.23 | -.14 | -.15 | -.17 | -.27 | -.31 | -.35 | .28 | .32 | .48 | .47 | -- |  |  |  |  |  |
| 31. Social Contact1 | 2846 | .17 | -.14 | .00 | -.09 | -.01 | .03 | .00 | .04 | .08 | .02 | .01 | -.02 | .04 | .02 | .05 | .10 | .02 | .01 | .02 | .04 | .04 | .06 | .01 | .00 | .02 | .04 | .06 | .05 | .11 | .07 | -- |  |  |  |  |
| 32. Social Contact2 | 2847 | .21 | -.22 | -.05 | -.16 | -.05 | .03 | .05 | .09 | .11 | .01 | -.01 | -.01 | .03 | .04 | .04 | .17 | .01 | .02 | .01 | .06 | .05 | .08 | .06 | .03 | .06 | .05 | .08 | .05 | .11 | .09 | .35 | -- |  |  |  |
| 33. Social Contact3 | 2772 | .17 | -.19 | -.03 | -.13 | -.03 | .03 | .00 | .07 | .11 | .01 | -.01 | .02 | .01 | .03 | .04 | .13 | -.02 | -.02 | .00 | .05 | .02 | .05 | .02 | .01 | .04 | .05 | .05 | .05 | .10 | .08 | .36 | .41 | -- |  |  |
| 34. Experience of Life Events1 | 2847 | .07 | -.14 | -.03 | -.12 | .01 | .04 | .06 | .03 | .07 | .07 | -.01 | .02 | .08 | .13 | .15 | .17 | .11 | .11 | .12 | .14 | .14 | .11 | .15 | .15 | .17 | -.04 | -.05 | -.04 | -.04 | -.06 | .06 | .08 | .05 | -- |  |
| 35. Experience of Life Events2 | 2847 | .05 | -.09 | -.05 | -.08 | -.01 | .01 | .09 | .01 | .05 | .05 | -.01 | .02 | .05 | .09 | .12 | .14 | .10 | .10 | .11 | .12 | .13 | .12 | .11 | .13 | .15 | -.04 | -.04 | -.06 | -.05 | -.05 | .05 | .04 | .06 | .42 | -- |

*Note.* Values in Bold indicated that the variables were too highly correlated to be used in the analysis.

## Appendix D

**Table 1.** Odds Ratio Table of Covariates Entered into the Model.

| Predictor | IFHD<br>[vs LRAS] |  | SIFD<br>[vs LRAS] |  | ISHB<br>[vs LRAS] |  | EBFD<br>[vs LRAS] |  |
| --- | --- | --- | --- | --- | --- | --- | --- | --- |
|  | OR | (95% CI) | OR | (95% CI) | OR | (95% CI) | OR | (95% CI) |
| Covariates measured pre-pandemic |  |  |  |  |  |  |  |  |
| Age group<sup>a</sup> |  |  |  |  |  |  |  |  |
| 18-20 | (ref) |  | (ref) |  | (ref) |  | (ref) |  |
| 49-50 | 2.38 | [1.51 , 3.75] | 1.96 | [1.27 , 3.02] | 5.07 | [3.66 , 7.04] | 8.05 | [5.21 , 12.43] |
| Gender<sup>a</sup> |  |  |  |  |  |  |  |  |
| Male | (ref) |  | (ref) |  | (ref) |  | (ref) |  |
| Female | 0.91 | [0.69 , 1.19] | 0.69 | [0.54 , 0.89] | 0.68 | [0.56 , 0.82] | 0.45 | [0.36 , 0.57] |
| Race<sup>a</sup> |  |  |  |  |  |  |  |  |
| Non-White Ethnic Group | (ref) |  | (ref) |  | (ref) |  | (ref) |  |
| White | 0.49 | [0.31 , 0.79] | 1.52 | [0.84 , 2.77] | 0.67 | [0.44 , 1.01] | 1.62 | [0.76 , 3.43] |
| Adverse childhood experiences |  |  |  |  |  |  |  |  |
| 0 ACEs | (ref) |  | (ref) |  | (ref) |  | (ref) |  |
| 1 ACE | 1.36 | [1.01 , 1.82] | 1.20 | [0.9 , 1.60] | 1.06 | [0.86 , 1.30] | 1.21 | [0.93 , 1.57] |
| 2 ACEs* | 1.18 | [0.84 , 1.67] | 1.77 | [1.29 , 2.41] | 1.08 | [0.85 , 1.38] | 1.04 | [0.76 , 1.43] |
| 3+ ACEs | 1.72 | [1.16 , 2.54] | 1.36 | [0.93 , 2.01] | 1.05 | [0.78 , 1.41] | 1.33 | [0.91 , 1.94] |
| Educational level |  |  |  |  |  |  |  |  |
| NVQ0 | 0.97 | [0.62 , 1.52] | 1.44 | [0.89 , 2.33] | 0.85 | [0.62 , 1.17] | 1.09 | [0.73 , 1.62] |
| NVQ1 | (ref) |  | (ref) |  | (ref) |  | (ref) |  |
| NVQ2 | 0.88 | [0.47 , 1.67] | 1.58 | [0.84 , 2.96] | 1.42 | [0.91 , 2.20] | 1.36 | [0.75 , 2.44] |
| NVQ3 | 1.43 | [0.74 , 2.74] | 1.78 | [0.91 , 3.48] | 1.23 | [0.75 , 2.01] | 1.86 | [0.99 , 3.48] |
| NVQ4 | 1.19 | [0.68 , 2.08] | 1.63 | [0.91 , 2.93] | 1.27 | [0.85 , 1.90] | 1.86 | [1.11 , 3.10] |
| NVQ5 | 1.14 | [0.62 , 2.09] | 1.40 | [0.74 , 2.62] | 0.74 | [0.46 , 1.16] | 1.42 | [0.77 , 2.59] |

| Covariates measured during T1 |  |  |  |  |  |  |  |  |
| --- | --- | --- | --- | --- | --- | --- | --- | --- |
| Mental health diagnosis |  |  |  |  |  |  |  |  |
| No | (ref) |  | (ref) |  | (ref) |  | (ref) |  |
| Yes | 0.80 | [0.51 , 1.24] | 1.04 | [0.70 , 1.54] | 1.15 | [0.83 , 1.60] | 0.58 | [0.37 , 0.91] |
| Psychological distress<sup>a</sup> |  |  |  |  |  |  |  |  |
| No | (ref) |  | (ref) |  | (ref) |  | (ref) |  |
| Yes | 1.25 | [0.84 , 1.85] | 1.57 | [1.07 , 2.29] | 1.67 | [1.22 , 2.29] | 2.47 | [1.68 , 3.64] |
| Above cut off score on PHQ-2GAD-2 |  |  |  |  |  |  |  |  |
| No | (ref) |  | (ref) |  | (ref) |  | (ref) |  |
| Yes | 0.89 | [0.59 , 1.34] | 1.11 | [0.75 , 1.64] | 0.82 | [0.59 , 1.14] | 0.82 | [0.52 , 1.28] |
| Instrumental support |  |  |  |  |  |  |  |  |
| No | (ref) |  | (ref) |  | (ref) |  | (ref) |  |
| Yes | 0.68 | [0.47 , 0.98] | 0.73 | [0.52 , 1.02] | 0.86 | [0.65 , 1.13] | 1.01 | [0.71 , 1.43] |
| Social Contact |  |  |  |  |  |  |  |  |
| No | (ref) |  | (ref) |  | (ref) |  | (ref) |  |
| Yes | 1.00 | [0.71 , 1.40] | 1.01 | [0.74 , 1.38] | 0.92 | [0.73 , 1.17] | 0.94 | [0.70 , 1.25] |
| Covariates measured during T2 |  |  |  |  |  |  |  |  |
| Experience of adverse life events<sup>a</sup> |  |  |  |  |  |  |  |  |
| No | (ref) |  | (ref) |  | (ref) |  | (ref) |  |
| Yes | 1.28 | [0.99 , 1.66] | 1.43 | [1.12 , 1.83] | 0.96 | [0.79 , 1.16] | 1.24 | [0.98 , 1.58] |
| Psychological distress |  |  |  |  |  |  |  |  |
| No | (ref) |  | (ref) |  | (ref) |  | (ref) |  |
| Yes | 1.53 | [1.03 , 2.26] | 1.41 | [0.96 , 2.07] | 1.18 | [0.86 , 1.61] | 1.41 | [0.95 , 2.10] |
| Above cut off score on PHQ-2GAD-2 |  |  |  |  |  |  |  |  |
| No | (ref) |  | (ref) |  | (ref) |  | (ref) |  |
| Yes | 0.90 | [0.59 , 1.38] | 1.07 | [0.73 , 1.58] | 0.89 | [0.64 , 1.23] | 0.71 | [0.45 , 1.13] |
| Loneliness score<sup>a</sup> | 1.27 | [1.09 , 1.48] | 1.09 | [0.94 , 1.26] | 1.20 | [1.07 , 1.34] | 0.91 | [0.78 , 1.06] |
| Instrumental support |  |  |  |  |  |  |  |  |
| No | (ref) |  | (ref) |  | (ref) |  | (ref) |  |
| Yes | 1.20 | [0.82 , 1.77] | 0.76 | [0.54 , 1.08] | 0.95 | [0.71 , 1.26] | 1.24 | [0.86 , 1.80] |
| Emotional support |  |  |  |  |  |  |  |  |
| No | (ref) |  | (ref) |  | (ref) |  | (ref) |  |
| Yes | 1.09 | [0.73 , 1.63] | 1.10 | [0.75 , 1.61] | 1.25 | [0.92 , 1.70] | 0.81 | [0.56 , 1.18] |
| Social Contact |  |  |  |  |  |  |  |  |
| No | (ref) |  | (ref) |  | (ref) |  | (ref) |  |
| Yes | 1.54 | [1.1 , 2.15] | 1.37 | [1.01 , 1.85] | 1.17 | [0.93 , 1.46] | 1.11 | [0.85 , 1.46] |
| Covariates measured during T3 |  |  |  |  |  |  |  |  |
| Mental Health Diagnosis |  |  |  |  |  |  |  |  |
| No | (ref) |  | (ref) |  | (ref) |  | (ref) |  |
| Yes | 1.14 | [0.76 , 1.70] | 1.35 | [0.93 , 1.96] | 1.32 | [0.97 , 1.79] | 1.99 | [1.36 , 2.91] |
| Psychological distress |  |  |  |  |  |  |  |  |
| No | (ref) |  | (ref) |  | (ref) |  | (ref) |  |
| Yes | 1.84 | [1.25 , 2.71] | 1.11 | [0.75 , 1.63] | 1.29 | [0.95 , 1.76] | 0.99 | [0.66 , 1.48] |
| Instrumental support |  |  |  |  |  |  |  |  |
| No | (ref) |  | (ref) |  | (ref) |  | (ref) |  |
| Yes | 1.06 | [0.70 , 1.61] | 0.88 | [0.60 , 1.29] | 1.00 | [0.74 , 1.36] | 0.91 | [0.62 , 1.35] |
| Social Contact |  |  |  |  |  |  |  |  |
| No | (ref) |  | (ref) |  | (ref) |  | (ref) |  |
| Yes | 0.87 | [0.64 , 1.18] | 0.76 | [0.57 , 1.00] | 0.98 | [0.79 , 1.22] | 0.76 | [0.59 , 0.99] |
| Experience of adverse life events<sup>a</sup> |  |  |  |  |  |  |  |  |
| No | (ref) |  | (ref) |  | (ref) |  | (ref) |  |
| Yes | 0.93 | [0.72 , 1.21] | 1.13 | [0.88 , 1.44] | 1.37 | [1.14 , 1.65] | 1.25 | [0.98 , 1.59] |
| Above cut off score on PHQ-2GAD-2 |  |  |  |  |  |  |  |  |
| No | (ref) |  | (ref) |  | (ref) |  | (ref) |  |
| Yes | 0.78 | [0.52 , 1.17] | 0.74 | [0.50 , 1.09] | 0.84 | [0.61 , 1.15] | 0.95 | [0.62 , 1.47] |
| Emotional support |  |  |  |  |  |  |  |  |
| No | (ref) |  | (ref) |  | (ref) |  | (ref) |  |
| Yes | 0.76 | [0.51 , 1.13] | 0.95 | [0.65 , 1.41] | 0.84 | [0.62 , 1.15] | 1.00 | [0.67 , 1.48] |

<sup>a</sup>Reflects significant covariates. All other covariates are not significant at $p < .05$

## Footnotes

1 We initially used a latent transition analysis to explore how people transition between subgroups, during the first nine months of the pandemic (Lanza et al., 2003). However, as explain in Appendix A, the decision was subsequently made to conduct a repeated measures latent class analysis (RMLCA).

## Notes

### Competing Interest Statement

The authors have declared no competing interest.

### Funding Statement

This study did not receive any funding.

### Author Declarations

The Ethics Committee of the University of Southampton gave ethical approval for this work (ERGO: 62740.A2).

## References

Acuff, S. F., Strickland, J. C., Tucker, J. A., & Murphy, J. G. (2022). Changes in alcohol use during COVID-19 and associations with contextual and individual difference variables: A systematic review and meta-analysis. Psychology of Addictive Behaviors, 36(1), 1–19. https://doi.org/10.1037/adb0000796.

Agresti, A. (2003). Categorical data analysis. John Wiley & Sons.

Akaike, H. (1974). A new look at the statistical model identification. IEEE Transactions on Automatic Control, 19(6), 716–723.

Al-Aly, Z, (2022, February 17). Mental health in people with covid-19 [Editorial]. BMJ. https://www.bmj.com/content/376/bmj.o415#:~:text=Altogether%2C%20the%20findings%20suggest%20that,opioid%20use%20disorders%20pre%2Dpandemic.

Aldwin, C. M. & Yancura, L. A. (2004). Coping. In C. Spielberger (Eds.), Encyclopaedia of Applied Psychology. (pp. 507–510). Academic Press.

Altena, E., Baglioni, C., Espie, C. A., Ellis, J., Gavriloff, D., Holzinger, B., Schlarb, A., Frase, L., Jernelöv, S., & Riemann, D. (2020). Dealing with sleep problems during home confinement due to the COVID-19 outbreak: Practical recommendations from a task force of the European CBT-I Academy. Journal of Sleep Research, 29(4), e13052. https://doi.org/10.1111/jsr.13052.

Appleton, S., Adams, R., Wilson, D., Taylor, A., Dal Grande, E., Chittleborough, C.,… & North West Adelaide Health (Cohort) Study team. (2004). The biomedically assessed cumulative burden of chronic disease risk factors on health-related quality of life in an urban population. Health Promotion Journal of Australia, 15(1), 55–62. https://doi.org/10.1071/HE04055.

Attard, R., Dingli, P., Doggen, C. J., Cassar, K., Farrugia, R., & Bezzina Wettinger, S. (2021). The impact of frequency, pattern, intensity, and type of alcohol consumption, and its combined effect with smoking on inflammation, lipid profile, and the risk of myocardial infarction. Journal of Public Health, 29, 611–624.

Atzl, V. M., Narayan, A. J., Rivera, L. M., & Lieberman, A. F. (2019). Adverse childhood experiences and prenatal mental health: Type of ACEs and age of maltreatment onset. Journal of Family Psychology, 33(3), 304. https://doi.org/10.1037/fam0000510.

Babor, T. J., Higgins-Biddle, J. C., Saunders, J. B., & Monteiro, M. G. (2001). AUDIT. The alcohol Use Disorders Test. Guidelines for Use in Primary Care. World Health Organization. Department of Mental Health and Substance Dependence.

Baguley, T. (2004). Understanding statistical power in the context of applied research. Applied Ergonomics, 35(2), 73–80. https://doi.org/10.1016/j.apergo.2004.01.002.

Bauer-Staeb, C., Davis, A., Smith, T., Wilsher, W., Betts, D., Eldridge, C., … & Button, K. S. (2021). The early impact of COVID-19 on primary care psychological therapy services: A descriptive time series of electronic healthcare records. EClinicalMedicine, 37, 100939. https://doi.org/10.1016/j.eclinm.2021.100939.

Bell, L. M., Smith, R., van de Venter, E. C., Shuttleworth, C., Wilson, K., & Lycett, D. (2021). COVID-19 stressors, wellbeing and health behaviours: a cross-sectional study. Journal of Public Health, 43(3), e453–e461.

Besedovsky, L., Lange, T., & Born, J. (2012). Sleep and immune function. European Journal of Physiology, 463(1), 121–137. https://doi.org/10.1007/s00424-011-1044.

Bevilacqua, L., Kelly, Y., Heilmann, A., Priest, N., & Lacey, R. E. (2021). Adverse childhood experiences and trajectories of internalizing, externalizing, and prosocial behaviors from childhood to adolescence. Child Abuse & Neglect, 112, 104890. https://doi.org/10.1016/j.chiabu.2020.104890

Blevins, C. E., Abrantes, A. M., & Stephens, R. S. (2016). Motivational pathways from antecedents of alcohol use to consequences: a structural model of using alcohol to cope with negative affect. The American journal of Drug and Alcohol abuse, 42(4), 395–403. https://doi.org/10.3109/00952990.2016.1141915.

Bowling A. (2005). Just one question: If one question works, why ask several?. Journal of Epidemiology and Community Health, 59(5), 342–345. https://doi.org/10.1136/jech.2004.021204

Bozdogan, H. (1987). Model selection and Akaike’s Information Criterion (AIC): The general theory and its analytical extensions. Psychometrika, 52(3), 345–370. https://doi.org/10.1007/BF02294361

Brookes, G. (2021). ‘Lose weight, save the NHS’: Discourses of obesity in press coverage of COVID-19. Critical Discourse Studies, 1–19.

Brown, S. P., Westbrook, R. A., & Challagalla, G. (2005). Good cope, bad cope: adaptive and maladaptive coping strategies following a critical negative work event. Journal of Applied Psychology, 90(4), 792.

Brown, S. P., Westbrook, R. A., & Challagalla, G. (2005). Good Cope, Bad Cope: Adaptive and Maladaptive Coping Strategies Following a Critical Negative Work Event. Journal of Applied Psychology, 90(4), 792–798. https://doi.org/10.1037/0021-9010.90.4.792.

Cairney, J., Kwan, M. Y., Veldhuizen, S., & Faulkner, G. E. (2014). Who uses exercise as a coping strategy for stress? Results from a national survey of Canadians. Journal of Physical Activity and Health, 11(5), 908–916.

Cameron-Blake, E., Tatlow, H., Wood, A., Hale, T., Kira, B., Petherick, A., & Phillips, T. (2020). Variation in the response to COVID-19 across the four nations of the United Kingdom. Blavatnik School of Government, University of Oxford.

Camilleri, G. M., Méjean, C., Kesse-Guyot, E., Andreeva, V. A., Bellisle, F., Hercberg, S., & Péneau, S. (2014). The associations between emotional eating and consumption of energy-dense snack foods are modified by sex and depressive symptomatology. The Journal of nutrition, 144(8), 1264–1273. https://doi.org/10.3945/jn.114.193177

Cardi, V., Leppanen, J., & Treasure, J. (2015). The effects of negative and positive mood induction on eating behaviour: A meta-analysis of laboratory studies in the healthy population and eating and weight disorders. Neuroscience and Biobehavioral Reviews, 57, 299–309. https://doi.org/10.1016/j.neubiorev.2015.08.011

Carvalho, J. P., & Hopko, D. R. (2011). Behavioral theory of depression: reinforcement as a mediating variable between avoidance and depression. Journal of Behavior Therapy and Experimental Psychiatry, 42(2), 154–162. https://doi.org/10.1016/j.jbtep.2010.10.001

Centre for Longitudinal Studies. (2010). User Guide to Analysing MCS Data Using SPSS. https://cls.ucl.ac.uk/wp-content/uploads/2017/06/Complex-Samples-in-SPSS.pdf

Chennaoui, M., Arnal, P. J., Sauvet, F., & Léger, D. (2015). Sleep and exercise: a reciprocal issue?. Sleep medicine reviews, 20, 59–72. https://doi.org/10.1016/j.smrv.2014.06.008

Cohen, J. (2013). Statistical power analysis for the behavioral sciences. Routledge.

Collins, L. M., & Lanza, S. T. (2010). Latent class and latent transition analysis: with applications in the social, behavioral, and health sciences (Vol. 718). John Wiley & Sons.

COVID-19 Mental Disorders Collaborators (2021). Global prevalence and burden of depressive and anxiety disorders in 204 countries and territories in 2020 due to the COVID-19 pandemic. Lancet (London, England), 398(10312), 1700–1712. https://doi.org/10.1016/S0140-6736(21)02143-7

Dempster, A. P., Laird, N. M., & Rubin, D. B. (1977). Maximum likelihood from incomplete data via the EM algorithm. Journal of the Royal Statistical Society, 39(1), 1–22

Dicken, S. J., Mitchell, J. J., Newberry Le Vay, J., Beard, E., Kale, D., Herbec, A., & Shahab, L. (2021). Impact of COVID-19 Pandemic on Weight and BMI among UK Adults: A Longitudinal Analysis of Data from the HEBECO Study. Nutrients, 13(9), 2911. https://doi.org/10.3390/nu13092911

Do, C. B., & Batzoglou, S. (2008). What is the expectation maximization algorithm?. Nature Biotechnology, 26(8), 897–899.

Dragioti, E., Li, H., Tsitsas, G., Lee, K. H., Choi, J., Kim, J., Choi, Y. J., Tsamakis, K., Estradé, A., Agorastos, A., Vancampfort, D., Tsiptsios, D., Thompson, T., Mosina, A., Vakadaris, G., Fusar-Poli, P., Carvalho, A. F., Correll, C. U., Han, Y. J., Park, S., … Solmi, M. (2022). A large-scale meta-analytic atlas of mental health problems prevalence during the COVID-19 early pandemic. Journal of Medical Virology, 94(5), 1935–1949. https://doi.org/10.1002/jmv.27549

Driscoll, K., & Pianta, R. C. (2011). Mothers’ and fathers’ perceptions of conflict and closeness in parent-child relationships during early childhood. Journal of Early Childhood and Infant Psychology, 7, 1–24.

Elliott, J., & Shepherd, P. (2006). Cohort profile: 1970 British birth cohort (BCS70). International Journal of Epidemiology, 35(4), 836–843.

Enders, C. K., & Bandalos, D. L. (2001). The relative performance of full information maximum likelihood estimation for missing data in structural equation models. Structural Equation Modeling, 8(3), 430–457. https://doi.org/10.1207/S15328007SEM0803_5

Eye, A., & Wiedermann, W. (2015). Person-Centered Analysis. In R. Scott, J. Lindsley & S. Kosslyn, Emerging Trends in the Social and Behavioral Sciences: Interdisciplinary Directions (1st ed., pp. 1–18). SAGE Publications. Retrieved 6 September 2022, from.

Feldman, C., & Anderson, R. (2013). Cigarette smoking and mechanisms of susceptibility to infections of the respiratory tract and other organ systems. The Journal of infection, 67(3), 169–184. https://doi.org/10.1016/j.jinf.2013.05.004

Felitti, V. J., Anda, R. F., Nordenberg, D., Williamson, D. F., Spitz, A. M., Edwards, V., Koss, M. P., & Marks, J. S. (1998). Relationship of childhood abuse and household dysfunction to many of the leading causes of death in adults. The Adverse Childhood Experiences (ACE) Study. American Journal of Preventive Medicine, 14(4), 245–258. https://doi.org/10.1016/s0749-3797(98)00017-8

Fitzpatrick, S. L., Coughlin, J. W., Appel, L. J., Tyson, C., Stevens, V. J., Jerome, G. J., Dalcin, A., Brantley, P. J., & Hill-Briggs, F. (2015). Application of Latent Class Analysis to Identify Behavioral Patterns of Response to Behavioral Lifestyle Interventions in Overweight and Obese Adults. International Journal of Behavioral Medicine, 22(4), 471–480. https://doi.org/10.1007/s12529-014-9446-y

Frayn, M., & Knäuper, B. (2018). Emotional eating and weight in adults: A review. Current Psychology: A Journal for Diverse Perspectives on Diverse Psychological Issues, 37(4), 924–933. https://doi.org/10.1007/s12144-017-9577-9

Goffe, L., Rushton, S., White, M., Adamson, A., & Adams, J. (2017) Relationship between mean daily energy intake and frequency of consumption of out-of-home meals in the UK National Diet and Nutrition Survey. International Journal of Behavioral Nutrition and Physical Activity, 14(1), 1–11

Goldmann, E., & Galea, S. (2014). Mental health consequences of disasters. Annual Review of Public Health, 35, 169–183. https://doi.org/10.1146/annurev-publhealth-032013-182435

Hirshkowitz, M., Whiton, K., Albert, S. M., Alessi, C., Bruni, O., DonCarlos, L., Hazen, N., Herman, J., Katz, E. S., Kheirandish-Gozal, L., Neubauer, D. N., O’Donnell, A. E., Ohayon, M., Peever, J., Rawding, R., Sachdeva, R. C., Setters, B., Vitiello, M. V., Ware, J. C., & Adams Hillard, P. J. (2015). National Sleep Foundation’s sleep time duration recommendations: methodology and results summary. Sleep Health, 1(1), 40–43. https://doi.org/10.1016/j.sleh.2014.12.010

HM Revenue & Customs (2020, July 15). Get a discount with the Eat Out to Help Out Scheme. https://www.gov.uk/guidance/get-a-discount-with-the-eat-out-to-help-out-scheme#:~:text=The%20Eat%20Out%20to%20Help%20Out%20Scheme%20closed%20on%2031%20August%202020.&text=Use%20the%20Eat%20Out%20to,between%203%20and%2031%20August

Holahan, C. J., Ragan, J. D., & Moos, R. H. (2004). Stress. In C. Spielberger (Eds.), Encylopedia of Applied Psychlogy. (pp. 485–492). Academic Press.

Hughes, J. M., Ulmer, C. S., Hastings, S. N., Gierisch, J. M., Mid-Atlantic VA MIRECC Workgroup, & Howard, M. O. (2018). Sleep, resilience, and psychological distress in United States military Veterans. Military Psychology, American Psychological Association, 30(5), 404–414. https://doi.org/10.1080/08995605.2018.147855

Institute for Government Analysis. (2020, February, 15). Timeline of UK coronavirus lockdowns, March 2020 to March 2021. https://www.instituteforgovernment.org.uk/sites/default/files/timeline-lockdown-web.pdf

Jackson, S. E., Garnett, C., Shahab, L., Oldham, M., & Brown, J. (2021). Association of the COVID-19 lockdown with smoking, drinking and attempts to quit in England: an analysis of 2019-20 data. Addiction (Abingdon, England), 116(5), 1233–1244. https://doi.org/10.1111/add.15295

Jahrami, H. A., Alhaj, O. A., Humood, A. M., Alenezi, A. F., Fekih-Romdhane, F., AlRasheed, M. M., Saif, Z. Q., Bragazzi, N. L., Pandi-Perumal, S. R., BaHammam, A. S., & Vitiello, M. V. (2022). Sleep disturbances during the COVID-19 pandemic: A systematic review, meta-analysis, and meta-regression. Sleep Medicine Reviews, 62, 101591. https://doi.org/10.1016/j.smrv.2022.10159

Joshi, H., & Fitzsimons, E. (2016). The Millennium Cohort Study: the making of a multi-purpose resource for social science and policy. Longitudinal and Life Course Studies, 7(4), 409–430.

Kantar Public. (2021). CLS COVID-19 Study – Waves 2 & 3. Centre for Longitudinal Studies (CLS). https://cls.ucl.ac.uk/wp-content/uploads/2017/02/CLS-COVID-19-Survey-Waves-2-and-3-technical-report.pdf

Karamanos, A., Stewart, K., Harding, S., Kelly, Y., & Lacey, R. E. (2022). Adverse childhood experiences and adolescent drug use in the UK: the moderating role of socioeconomic position and ethnicity. SSM-Population Health, 19, 101142.

Kass, L., Desai, T., Sullivan, K., Muniz, D., & Wells, A. (2021). Changes to Physical Activity, Sitting Time, Eating Behaviours and Barriers to Exercise during the First COVID-19 ‘Lockdown’ in an English Cohort. International Journal of Environmental Research and Public Health, 18(19), 10025. https://doi.org/10.3390/ijerph181910025

Kemp, E., Bui, M. Y., & Grier, S. (2013). When food is more than nutrition: Understanding emotional eating and overconsumption. Journal of Consumer Behaviour, 12(3), 204–213. https://doi.org/10.1002/cb.1413

Kessler, R. C., Barker, P. R., Colpe, L. J., Epstein, J. F., Gfroerer, J. C., Hiripi, E., Howes, M. J., Normand, S.-L. T., Manderscheid, R. W., Walters, E. E., & Zaslavsky, A. M. (2003). Screening for serious mental illness in the general population. Archives of General Psychiatry, 60(2), 184–189. https://doi.org/10.1001/archpsyc.60.2.184

Khan, A., Chien, C. W., & Burton, N. W. (2014). A new look at the construct validity of the K6 using Rasch analysis. International journal of methods in psychiatric research, 23(1), 1–8. https://doi.org/10.1002/mpr.1431

Khantzian, E. J. (2003). The self-medication hypothesis revisited: The dually diagnosed patient. Primary Psychiatry, 10(9), 47–54.

Kilian, C., Rehm, J., Allebeck, P., Braddick, F., Gual, A., Barták, M., Bloomfield, K., Gil, A., Neufeld, M., O’Donnell, A., Petruželka, B., Rogalewicz, V., Schulte, B., Manthey, J., & European Study Group on Alcohol Use and COVID-19 (2021). Alcohol consumption during the COVID-19 pandemic in Europe: a large-scale cross-sectional study in 21 countries. Addiction, 116(12), 3369–3380. https://doi.org/10.1111/add.15530

Kroenke, K., Spitzer, R. L., & Williams, J. B. (2003). The Patient Health Questionnaire-2: validity of a two-item depression screener. Medical Care, 41(11), 1284–1292. https://doi.org/10.1097/01.MLR.0000093487.78664.3C

Kroenke, K., Spitzer, R. L., Williams, J. B., Monahan, P. O., & Löwe, B. (2007). Anxiety disorders in primary care: prevalence, impairment, comorbidity, and detection. Annals of Internal Medicine, 146(5), 317–325. https://doi.org/10.7326/0003-4819-146-5-200703060-00004

Lacey, R. E., Bartley, M., Pikhart, H., Stafford, M., & Cable, N. (2014). Parental separation and adult psychological distress: an investigation of material and relational mechanisms. BMC Public Health, 14, 272. https://doi.org/10.1186/1471-2458-14-272

Langer, E. J. (1977). The psychology of chance. Journal for the Theory of Social Behaviour, 7(2), 185–207. https://doi.org/10.1111/j.1468-5914.1977.tb00384.x

Lanza, S. T., & Collins, L. M. (2006). A mixture model of discontinuous development in heavy drinking from ages 18 to 30: the role of college enrollment. Journal of Studies on Alcohol, 67(4), 552–561. https://doi.org/10.15288/jsa.2006.67.552

Lanza, S. T., Dziak, J. J., Huang, L., Wagner, A., & Collins, L. M. (2015). Proc LCA & Proc LTA users’ guide (Version 1.3. 2). University Park: The Methodology Center, Penn State.

Lanza, S. T., Flaherty, B. P., & Collins, L. M. (2003). Latent class and latent transition analysis. In J. A. Schinka & W. F. Velicer (Eds.), Handbook of psychology: Research Methods in Psychology, Vol. 2, pp. 663–685). John Wiley & Sons Inc. https://doi.org/10.1002/0471264385.wei0226

Lanza, S. T., & Rhoades, B. L. (2013). Latent class analysis: an alternative perspective on subgroup analysis in prevention and treatment. Prevention science, 14(2), 157–168. https://doi.org/10.1007/s11121-011-0201-1

Lazarus, R., & Folkman, S. (1984). Stress, appraisal, and coping. Springer.

Lewinsohn, P. M. (1974). A behavioral approach to depression. Essential Papers on Depression, 150–172.

Löwe, B., Wahl, I., Rose, M., Spitzer, C., Glaesmer, H., Wingenfeld, K., Schneider, A., & Brähler, E. (2010). A 4-item measure of depression and anxiety: validation and standardization of the Patient Health Questionnaire-4 (PHQ-4) in the general population. Journal of Affective Disorders, 122(1-2), 86–95. https://doi.org/10.1016/j.jad.2009.06.019

Macht M. (2008). How emotions affect eating: a five-way model. Appetite, 50(1), 1–11. https://doi.org/10.1016/j.appet.2007.07.002

MacKillop, J., Amlung, M. T., Wier, L. M., David, S. P., Ray, L. A., Bickel, W. K., & Sweet, L. H. (2012). The neuroeconomics of nicotine dependence: a preliminary functional magnetic resonance imaging study of delay discounting of monetary and cigarette rewards in smokers. Psychiatry Research, 202(1), 20–29. https://doi.org/10.1016/j.pscychresns.2011.10.003

Mahmood, S., Li, Y., & Hynes, M. (2022). Adverse Childhood Experiences and Obesity: A One-to-One Correlation?. Clinical Child Psychology and Psychiatry, 13591045221119001. https://doi.org/10.1177/13591045221119001

Mandelkorn, U., Genzer, S., Choshen-Hillel, S., Reiter, J., Meira E Cruz, M., Hochner, H., Kheirandish-Gozal, L., Gozal, D., & Gileles-Hillel, A. (2021). Escalation of sleep disturbances amid the COVID-19 pandemic: a cross-sectional international study. Journal of Clinical Sleep Medicine, 17(1), 45–53. https://doi.org/10.5664/jcsm.8800

Maric, V., Mishra, J., & Ramanathan, D. S. (2021). Using Mind-Body Medicine to Reduce the Long-Term Health Impacts of COVID-Specific Chronic Stress. Frontiers in Psychiatry, 12, 585952. https://doi.org/10.3389/fpsyt.2021.585952

Marryat, L., & Frank, J. (2019). Factors associated with adverse childhood experiences in Scottish children: a prospective cohort study. BMJ Paediatrics Open, 3(1), e000340. https://doi.org/10.1136/bmjpo-2018-000340

McAtamney, K., Mantzios, M., Egan, H., & Wallis, D. J. (2021). Emotional eating during COVID-19 in the United Kingdom: Exploring the roles of alexithymia and emotion dysregulation. Appetite, 161, 105120. https://doi.org/10.1016/j.appet.2021.105120

McCrory, E. J., Gerin, M. I., & Viding, E. (2017). Annual Research Review: Childhood maltreatment, latent vulnerability and the shift to preventative psychiatry - the contribution of functional brain imaging. Journal of Child Psychology and Psychiatry, and Allied Disciplines, 58(4), 338–357. https://doi.org/10.1111/jcpp.1271

McCrory, E. J., & Viding, E. (2015). The theory of latent vulnerability: Reconceptualizing the link between childhood maltreatment and psychiatric disorder. Development and Psychopathology, 27(2), 493–505. https://doi.org/10.1017/S0954579415000115

McKee, S. A., Sinha, R., Weinberger, A. H., Sofuoglu, M., Harrison, E. L., Lavery, M., & Wanzer, J. (2011). Stress decreases the ability to resist smoking and potentiates smoking intensity and reward. Journal of Psychopharmacology, 25(4), 490–502. https://doi.org/10.1177/0269881110376694

McLaughlin, K. A., Peverill, M., Gold, A. L., Alves, S., & Sheridan, M. A. (2015). Child Maltreatment and Neural Systems Underlying Emotion Regulation. Journal of the American Academy of Child and Adolescent Psychiatry, 54(9), 753–762. https://doi.org/10.1016/j.jaac.2015.06.01

McPhie, M. L., Weiss, J. A., & Wekerle, C. (2014). Psychological distress as a mediator of the relationship between childhood maltreatment and sleep quality in adolescence: results from the Maltreatment and Adolescent Pathways (MAP) Longitudinal Study. Child abuse & Neglect, 38(12), 2044–2052. https://doi.org/10.1016/j.chiabu.2014.07.009

Miller, W. C., Koceja, D. M., & Hamilton, E. J. (1997). A meta-analysis of the past 25 years of weight loss research using diet, exercise or diet plus exercise intervention. International Journal of Obesity, 21(10), 941–947.

Morelli, S. A., Lee, I. A., Arnn, M. E., & Zaki, J. (2015). Emotional and instrumental support provision interact to predict well-being. Emotion, 15(4), 484–493. https://doi.org/10.1037/emo000008

Morgan, Z., Brugha, T., Fryers, T., & Stewart-Brown, S. (2012). The effects of parent-child relationships on later life mental health status in two national birth cohorts. Social Psychiatry and Psychiatric Epidemiology, 47(11), 1707–1715. https://doi.org/10.1007/s00127-012-0481-1

Mostafa, T., & Wiggins, D. (2014). Handling attrition and non-response in the 1970 British Cohort Study. https://cls.ucl.ac.uk/wp-content/uploads/2017/04/CLS-WP-2014-2.pdf

National Institute on Alcohol Abuse and Alcoholism (n.d.). Drinking Levels Defined. https://www.niaaa.nih.gov/alcohol-health/overview-alcohol-consumption/moderate-binge-drinking

National Health Service. (2018). Alcohol units. https://www.nhs.uk/live-well/alcohol-advice/calculating-alcohol-units/#:~:text=men%20and%20women%20are%20advised,drink%2Dfree%20days%20each%20week.

National Health Service (2019, January). The NHS Long Term Plan. National Health Service. https://www.longtermplan.nhs.uk/wp-content/uploads/2019/01/nhs-long-term-plan-june-2019.pdf.

National Health Service. (2020, May). Statistics on Obesity, Physical Activity and Diet, England, 2020. Official Statistics, National statistics. https://www.nhs.uk/live-well/alcohol-advice/calculating-alcohol-units/#:~:text=men%20and%20women%20are%20advised,drink%2Dfree%20days%20each%20week.

National Mental Health Intelligence Network. (2022). COVID-19 mental health and wellbeing surveillance: report. Office for Health Improvement and Disparities. https://www.gov.uk/government/publications/covid-19-mental-health-and-wellbeing-surveillance-report.

Nylund-Gibson, K., & Choi, A. Y. (2018). Ten frequently asked questions about latent class analysis. Translational Issues in Psychological Science, 4(4), 440–461. https://doi.org/10.1037/tps0000176

Office for National Statistics. (2020). Adult smoking habits in the UK: 2019. https://www.ons.gov.uk/peoplepopulationandcommunity/healthandsocialcare/healthandlifeexpectancies/bulletins/adultsmokinghabitsingreatbritain/2019

Office for National Statistics. (2021). Alcohol-specific deaths in the UK: registered in 2019. https://www.ons.gov.uk/peoplepopulationandcommunity/healthandsocialcare/causesofdeath/bulletins/alcoholrelateddeathsintheunitedkingdom/registeredin2019

Ogueji, I. A., Okoloba, M. M., & Demoko Ceccaldi, B. M. (2021). Coping strategies of individuals in the United Kingdom during the COVID-19 pandemic. Current Psychology, 1–7. https://doi.org/10.1007/s12144-020-01318-7

Phelan, C. H., Love, G. D., Ryff, C. D., Brown, R. L., & Heidrich, S. M. (2010). Psychosocial predictors of changing sleep patterns in aging women: a multiple pathway approach. Psychology and Aging, 25(4), 858.

Prochaska, J. O. (2020). Transtheoretical model of behavior change. Encyclopedia of Behavioral Medicine, 2266–2270.

Public Health England (2021). Monitoring alcohol consumption and harm during the COVID-19 pandemic. https://assets.publishing.service.gov.uk/government/uploads/system/uploads/attachment_data/file/1002627/Alcohol_and_COVID_report.pdf

Qiu, L., Chan, S. H. M., & Chan, D. (2018). Big data in social and psychological science: theoretical and methodological issues. Journal of Computational Social Science, 1(1), 59–66.

Rabow, J., & Watts, R. K. (1983). The role of alcohol availability in alcohol consumption and alcohol problems. Recent developments in Alcoholism, 1, 285–302. https://doi.org/10.1007/978-1-4613-3617-4_17

Rodgers B. (1996). Reported parental behaviour and adult affective symptoms. 1. Associations and moderating factors. Psychological medicine, 26(1), 51–61. https://doi.org/10.1017/s0033291700033717

Rodgers, B., Pickles, A., Power, C., Collishaw, S., & Maughan, B. (1999). Validity of the Malaise Inventory in general population samples. Social psychiatry and psychiatric epidemiology, 34(6), 333–341. https://doi.org/10.1007/s001270050153

Rodrigues, H., Valentin, D., Franco-Luesma, E., Rakotosamimanana, V. R., Gomez-Corona, C., Saldaña, E., & Sáenz-Navajas, M. P. (2022). How has COVID-19, lockdown and social distancing changed alcohol drinking patterns? A cross-cultural perspective between britons and spaniards. Food Quality and Preference, 95, 104344.

Rutter, M., Tizard, J., & Whitmore, K. (1981). Education, health, and behaviour. R.E. Krieger Pub. Co.

Russell, A. M., & Barry, A. E. (2021). Psychometric Properties of the AUDIT-C within an Amazon Mechanical Turk Sample. American Journal of Health Behavior, 45(4), 695–700. https://doi.org/10.5993/AJHB.45.4.8

Russell, D. W. (1996). UCLA Loneliness Scale (Version 3): Reliability, validity, and factor structure. Journal of Personality Assessment, 66(1), 20–40. https://doi.org/10.1207/s15327752jpa6601_2

Sacco, K., George, T., Head, C., Vessicchio, J., Easton, C., & Prigerson, H. (2007). Adverse Childhood Experiences, Smoking and Mental Illness in Adulthood: A Preliminary Study. Annals of Clinical Psychiatry, 19(2), 89–97. https://doi.org/10.1080/10401230701334762

Salazar-Fernández, C., Mawditt, C., Palet, D., Haeger, P. A., & Román Mella, F. (2022). Changes in the clustering of health-related behaviors during the COVID-19 pandemic: examining predictors using latent transition analysis. BMC public health, 22(1), 1446. https://doi.org/10.1186/s12889-022-13854-x

SAS Institute Inc. (2016). SAS/SHARE® 9.4: User’s guide (2nd ed.). http://documentation.sas.com/api/docsets/shrref/9.4/content/shrref.pdf?locale=en#nameddest=bookinfo

Schoenborn, C., & Adams, P. (2008). Sleep Duration as a Correlate of Smoking, Alcohol Use, Leisure-Time Physical Inactivity, and Obesity Among Adults: United States, 2004-2006. Division of Health Interview Statistics. https://www.cdc.gov/nchs/data/hestat/sleep04-06/sleep04-06.pdf

Schwarz, G. (1978). Estimating the dimension of a model. The Annals of Statistics, 461–464.

Sclove, S. L. (1987). Application of model-selection criteria to some problems in multivariate analysis. Psychometrika, 52(3), 333–343. https://doi.org/10.1007/BF02294360

Shen, W., Long, L. M., Shih, C. H., & Ludy, M. J. (2020). A Humanities-Based Explanation for the Effects of Emotional Eating and Perceived Stress on Food Choice Motives during the COVID-19 Pandemic. Nutrients, 12(9), 2712. https://doi.org/10.3390/nu12092712

Soper, D.S. (2022). A-priori Sample Size Calculator for Structural Equation Models [Software]. Available from https://www.danielsoper.com/statcalc

Spring, B., Moller, A. C., & Coons, M. J. (2012). Multiple health behaviours: overview and implications. Journal of Public Health, 34 Suppl 1(Suppl 1), i3–i10. https://doi.org/10.1093/pubmed/fdr111

Sullivan, A., Brown, M., Hamer, M., & Ploubidis, G. B. (2022). Cohort Profile Update: The 1970 British Cohort Study (BCS70). International Journal of Epidemiology, dyac148. https://doi.org/10.1093/ije/dyac148

Sultana, A., Tasnim, S., Hossain, M. M., Bhattacharya, S., & Purohit, N. (2021). Digital screen time during the COVID-19 pandemic: a public health concern. F1000Research, 10(81), 81.

Szkody, E., Stearns, M., Stanhope, L., & McKinney, C. (2021). Stress-buffering role of social support during COVID-19. Family Process, 60(3), 1002–1015.

Tobin, R. M., & Graziano, W. G. (2010). Delay of gratification: A review of fifty years of regulation research. Handbook of Personality and Self-Regulation, 47–63.

Trepte, S., Dienlin, T., & Reinecke, L. (2015). Influence of social support received in online and offline contexts on satisfaction with social support and satisfaction with life: A longitudinal study. Media Psychology, 18(1), 74–105. https://doi.org/10.1080/15213269.2013.838904

University College London, UCL Institute of Education, Centre for Longitudinal Studies. (2021). COVID-19 Survey in Five National Longitudinal Cohort Studies: Millennium Cohort Study, Next Steps, 1970 British Cohort Study and 1958 National Child Development Study, 2020-2021. [data collection]. 3rd Edition. UK Data Service. SN: 8658, http://doi.org/10.5255/UKDA-SN-8658-3

Vandevijvere, S., Chow, C. C., Hall, K. D., Umali, E., & Swinburn, B. A. (2015). Increased food energy supply as a major driver of the obesity epidemic: a global analysis. Bulletin of the World Health Organization, 93(7), 446–456. https://doi.org/10.2471/BLT.14.150565

Vandekerckhove, M., & Wang, Y. L. (2017). Emotion, emotion regulation and sleep: An intimate relationship. AIMS Neuroscience, 5(1), 1–17. https://doi.org/10.3934/Neuroscience.2018.1.

Viertiö, S., Kiviruusu, O., Piirtola, M., Kaprio, J., Korhonen, T., Marttunen, M., & Suvisaari, J. (2021). Factors contributing to psychological distress in the working population, with a special reference to gender difference. BMC public health, 21(1), 1–17.

Villadsen, A., Patalay, P., & Bann, D. (2021). Mental health in relation to changes in sleep, exercise, alcohol and diet during the COVID-19 pandemic: examination of four UK cohort studies. Psychological Medicine, 1–10. https://doi.org/10.1017/S0033291721004657

Wang, M.-C., Deng, Q., Bi, X., Ye, H., & Yang, W. (2017). Performance of the entropy as an index of classification accuracy in latent profile analysis: A Monte Carlo simulation study. Acta Psychologica Sinica, 49(11), 1473–1482. https://doi.org/10.3724/SP.J.1041.2017.01473

Weller, B. E., Bowen, N. K., & Faubert, S. J. (2020). Latent Class Analysis: A Guide to Best Practice. Journal of Black Psychology, 46(4), 287–311. https://doi.org/10.1177/0095798420930932

Westland, J.C. (2010). Lower bounds on sample size in structural equation modelling. Electronic Commerce Research and Applications, 9(6), 476–487.

Whitson, J. A., & Galinsky, A. D. (2008). Lacking control increases illusory pattern perception. Science, 322(5898), 115–117. https://doi.org/10.1126/science.1159845

Wiss, D. A., & Brewerton, T. D. (2020). Adverse Childhood Experiences and Adult Obesity: A Systematic Review of Plausible Mechanisms and Meta-Analysis of Cross-Sectional Studies. Physiology & Behavior, 223, 112964. https://doi.org/10.1016/j.physbeh.2020.112964

Wood, N., Stafford, M., & O’Neill, D. (2019). CLOSER work package 9: Harmonised childhood environment and adult wellbeing measures user guide. CLOSER. https://doc.ukdataservice.ac.uk/doc/8553/mrdoc/pdf/closer_wp9_userguide_202203revision.pdf.

World Health Organisation. (2010). A healthy lifestyle – WHO recommendations. https://www.who.int/europe/news-room/fact-sheets/item/a-healthy-lifestyle---who-recommendations

World Health Organisation. (2022). Tobacco. https://www.who.int/news-room/fact-sheets/detail/tobacco

Wright, M. O. D., Crawford, E., & Del Castillo, D. (2009). Childhood emotional maltreatment and later psychological distress among college students: The mediating role of maladaptive schemas. Child Abuse & Neglect, 33(1), 59–68.

Wulfert, E., Block, J. A., Santa Ana, E., Rodriguez, M. L., & Colsman, M. (2002). Delay of gratification: Impulsive choices and problem behaviors in early and late adolescence. Journal of Personality, 70(4), 533–552.

Yoo, J. E., Shin, D. W., Han, K., Kim, D., Jeong, S. M., Koo, H. Y., Yu, S. J., Park, J., & Choi, K. S. (2021). Association of the Frequency and Quantity of Alcohol Consumption With Gastrointestinal Cancer. JAMA Network Open, 4(8), e2120382. https://doi.org/10.1001/jamanetworkopen.2021.20382

Zarse, E. M., Neff, M. R., Yoder, R., Hulvershorn, L., Chambers, J. E., & Chambers, R. A. (2019). The adverse childhood experiences questionnaire: Two decades of research on childhood trauma as a primary cause of adult mental illness, addiction, and medical diseases. Cogent Medicine, 6(1), 1581447.

Zhang, C., Xiao, S., Lin, H., Shi, L., Zheng, X., Xue, Y., Dong, F., Zhang, J., & Xue, B. (2022). The association between sleep quality and psychological distress among older Chinese adults: a moderated mediation model. BMC geriatrics, 22(1), 35. https://doi.org/10.1186/s12877-021-02711-y

